# Residential exposure to green and blue spaces over childhood and cardiometabolic health outcomes: The Generation XXI birth cohort

**DOI:** 10.1101/2024.10.23.24315979

**Authors:** Berta Valente, B. Araújo, Rita Pereira, Ana Isabel Ribeiro, Henrique Barros, Susana Santos

## Abstract

**Background:** Evidence on the effects of exposure to green and blue spaces on childhood cardiometabolic health is inconsistent, limited and mostly cross-sectional.

**Objectives:** To assess the associations of exposure to green and blue spaces, at birth, 4, 7, and 10 years (to identify vulnerable periods of exposure) and as longitudinal trajectories (to identify the longitudinal effect over time), with cardiometabolic health outcomes at 10 years.

**Methods:** Participants are from Generation XXI, a population-based birth cohort from Porto Metropolitan Area, Portugal (n=4669). Residential normalized difference vegetation index (NDVI) and Euclidian distance to the nearest urban green and blue space were assessed at birth, 4, 7 and 10 years using geographic information systems and standardized by dividing the observed value by the standard deviation. Longitudinal trajectories of exposures from birth to 10 years were derived using latent class mixed models. At 10 years, we measured body mass index, fat mass index and android-to-gynoid fat ratio, blood pressure, and metabolic outcomes. We defined overweight/obesity by the World Health Organization, high blood pressure by the American Academy of Pediatrics and metabolic syndrome by the IDEFICS study.

**Results:** No significant associations were observed between natural spaces exposure and body mass index, body fat content and distribution. We found an inverse association between distance to nearest blue space at birth and systolic blood pressure z-scores, and a positive association between distance to nearest green space at 7 and 10 years and metabolic syndrome score (p-values<0.05). Also, compared to children in the high stable trajectory of NDVI500m, those in the descending trajectory of NDVI500m presented a lower diastolic blood pressure z-score and metabolic syndrome score (p-values<0.05). However, after multiple testing correction, all associations lost statistical significance.

**Discussion:** This study did not find robust associations between the exposure to natural spaces over key developmental periods and cardiometabolic health.

## Introduction

More than half of the world’s population is projected to live in urban areas by 2050 ^1^, compromising the availability, accessibility, and quality of natural outdoor environments, such as green and blue spaces ^2^. These natural spaces have been associated with salutogenic effects ^3,4,5^, promoting physical activity opportunities^6,7^, psychological restoration, namely alleviation of stress, anxiety and depressive moods, as well as improved overall psychological well-being ^8,9,7,10^, reducing exposure to air pollution, and noise, lessening the urban heat island effect^7^, and facilitating social interactions^11,7^. While exposure to urban green and blue spaces has been shown to be protective in adulthood against cardiometabolic, respiratory, and neurodevelopmental morbidity and mortality ^3,12^, evidence in childhood remains limited. Understanding the effects of urban green and blue spaces on childhood health is critical, given that over 1 billion children worldwide currently live and grow in urban areas ^13^. This is also particularly relevant considering that childhood is a well-recognized period of high susceptibility to environmental damage with life-long consequences^14^.

Overweight/obesity and high blood pressure are major risk factors for non-communicable chronic diseases (NCDs), whose global prevalence in childhood is estimated to be up to 18% ^15^ and 9.7% ^16^, respectively. When co-occurring, alongside with dyslipidemia and hyperglycemia, these conditions can culminate in metabolic syndrome, which accounts for a global prevalence in children and adolescents of 3.3% ^17^ – further increasing the risk of cardiovascular disease in adulthood ^18^. Previous studies on the influence of exposure to green and blue spaces on overweight/obesity and high blood pressure in childhood show mixed results^19–39^. While the proximity to blue spaces has been associated with lower odds of childhood overweight and obesity^19,20^, studies on green spaces exposure have reported protective^21–24,36–39^ but also adverse^40,41^ or no^26–29^ effects on obesity in childhood. Some previous studies reported that greater exposure to greenness around children’s schools^30–33^ and residences^34^ was associated with lower blood pressure. Conversely, a study within the HELIX, a European consortium of six birth cohorts, found no association of green and blue spaces with paediatric blood pressure^35^. A recent individual participant data meta-analysis comprising 10 European cohorts found no association between green space and body mass index (BMI) and blood pressure during childhood^42^. To the best of our knowledge, no study has yet explored the influence of urban natural spaces on childhood body fat content and distribution, which is a better predictor of health risk than BMI, as well as on metabolic syndrome.

The existing studies differ in the methodological definition of natural space exposure. While those examining blue spaces focus on coastal distance^19,20^, studies on urban green spaces use a variety of exposure measures, such as greenness (Normalized Difference Vegetation Index)^22,25,26,30–34,38,39,43^, street tree density^23,32^, park access^29,36^, area^23,25^ and proximity^22,24,26,36^, as well as the percentage of urban green spaces^21,39^ and time spent in green spaces^28^, using them either in isolation or in combination. Additionally, most of the mentioned studies are cross-sectional^19,20,22–32,34,36,38^ and do not explore the potential cumulative effects and the time-variability of green and blue space exposures^44^. Therefore, adopting a life-course approach and assessing exposure to green and blue spaces over childhood will not only account for cumulative effects, but may also identify periods of vulnerability during early-life ^45^.

We aimed to investigate the associations of exposure to green and blue spaces at birth, 4, 7 and 10 years and as trajectories from birth until 10 years, with body mass index, body fat content and fat distribution, blood pressure and metabolic syndrome in children aged 10 years. We specifically aimed to identify vulnerable periods of exposure and the longitudinal effect over time. We hypothesized that a greater exposure to residential green and blue spaces from birth through late childhood is associated with improved cardiometabolic health at 10 years.

## Methods

### Study participants

This study is embedded within the Generation XXI (G21), a birth cohort from Porto Metropolitan Area, Portugal. G21 comprises 8495 mothers and 8647 newborns recruited between April 2005 and August 2006 in all five existing public maternity units. During the hospital stay, mothers who delivered live births with at least 24 gestational weeks were invited to participate in the study, of whom 92% agreed. Participants were re-evaluated at 4, 7, and 10 years of age (86%, 80%, and 74% participation rates, respectively). More cohort details were published elsewhere ^46,47^. In this study, we included singletons with complete information on exposure to urban natural spaces at birth, 4, 7, and 10 years (n=7549). From those, we excluded children without any information on cardiometabolic outcomes at 10 years and on covariates. Thus, 4669 children were included in the analysis (Figure S1).

All phases of the study complied with the Ethical Principles for Medical Research Involving Human Subjects expressed in the Declaration of Helsinki ^48^. The baseline and follow-up evaluations until 10 years were approved by the University of Porto Medical School/ S. João Hospital Centre Ethics Committee. At baseline and follow-up evaluations, all procedures were explained to participants, and an informed consent was signed by one of the parents or legal guardians. The baseline evaluation was additionally approved by the Data Protection National Commission and the study follows the present EU General Data Protection Regulation under close supervision of the Data Protection Office of ISPUP.

### Study area

Porto Metropolitan Area is the second largest metropolitan area of the country with almost 1.3 million inhabitants ^49^. It is in the northwest region Continental Portugal and is limited by the Atlantic Coast and extends along the Douro River estuary. Although Porto Metropolitan Area is constituted by 17 municipalities spread over almost 2.040 km^2^, 6 of them comprise nearly 85% of its population (N = 1,112,555 inhabitants) - Porto, Matosinhos, Maia, Vila Nova de Gaia, Gondomar, and Valongo^49^. Over the years, these municipalities experienced urbanization at different rates^50^. Porto, for example, turned from a “green city” with 76% of green areas in 1892 to a much more “grey city” with 29% of green areas remaining by 2000^51^.

### Urban natural spaces at birth, 4, 7 and 10 years

#### Greenness

Surrounding greenness was estimated using the normalized difference vegetation index (NDVI), which represents the density of vegetation, within 100m, 250m, and 500m of the child’s residence. Different buffers were used to cover immediate and more distant areas of exposure ^52^ NDVI was calculated based on land surface reflectance of visible red and near-infrared wavelengths. The values range from −1 (water) to 0 (rock, sand, and snow) and 1 (photosynthetically active and healthy vegetation), with higher positive values indicating denser green vegetation. In this study, negative NDVI values were omitted from the calculation. Only images with ≤ 5% of cloud coverage from Landsat 5 and 8 (spatial resolution: 30 m) during the spring/summer period (peak of vegetation) of the assessment years (2005/2006, 2009/2011, 2012/2014, and 2015/2017) were used, as previously described in G21 cohort studies ^53,54^. ArcMap 10.5 was used to process satellite images, and QGIS 3.8 was used to extract the average NDVI.

#### Residential distance to the nearest urban green space

The Euclidean distance (in hectometres) from the child’s residential address to the nearest urban green space (UGS) was measured during the G21 cohort evaluations. UGS referred to all public and free-to-access UGS in the Porto Metropolitan Area (n=662), regardless of size, location, or specific characteristics, namely public parks, gardens, and other green spaces managed by city councils of Porto Metropolitan Area or privately owned, but freely accessible. Cartography was obtained from digital maps provided by the city halls of the Porto Metropolitan Area. When UGS were fenced, the distance to the entrance was used; otherwise, the distance to the nearest boundary was considered. ArcGIS version 10.5 and the Network Analyst extension were used for calculations, applying an updated street network dataset provided by the Environmental Systems Research Institute.

#### Residential distance to the nearest urban blue space

The Euclidean distance (in hectometres) from the child’s residential address to the nearest river or sea was measured during the G21 cohort evaluations, using the Portuguese Water Atlas (from the Portuguese “*Atlas da Água*”). The Portuguese Water Atlas was developed by the National Water Resources Information System (SNIRH) under the responsibility of the Portuguese Environment Agency. The SNIRH provides information on hydrometeorological variables and water quality (surface, groundwater, and coastal) through the network of water resource monitoring stations in Portugal. In this study, other types of blue spaces such as fountains, ponds and lakes were not included due to their small size.

### Cardiometabolic outcomes at 10 years

#### Body mass index, body fat content and distribution

Height was measured using a stadiometer (SECA 206 Hamburg, Germany®) to the nearest 0.1 cm, and weight with a digital scale (TANITA® model TBF 300) to the nearest 0.1 kg, while the child stood barefooted in underwear. Body mass index (BMI) was calculated as weight (kg)/height (m)^2^ and then converted into standard deviation (SD) scores (BMI *z*-scores), adjusted for age and sex using the World Health Organization (WHO) growth reference values ^55^. The WHO cut-offs were applied, and the following BMI z-scores categories were defined: <−2 SDs for underweight, −2 ≤ SD ≤ 1 for normal weight, 1 > SD ≤ 2 for overweight, and >2 SDs for obesity ^55^. For models focused on the risk of overweight/obesity, children with underweight were excluded, and those with overweight and obesity were combined.

We measured total body fat mass, and android and gynoid fat mass by whole-body dual-energy X-ray absorptiometry (DXA) scans using a Hologic Discovery QDR 4500W device (Hologic Inc., Bedford, Massachusetts) and following the standard manufacturer’s protocol, with the child in underwear and with an emptied bladder^56^. We divided total fat mass by height^4^ in order to obtain a fat mass index (FMI) uncorrelated with height after estimating the optimal adjustment through log-log regression analyses^57^. We also calculated android-to-gynoid fat ratio dividing android by gynoid fat mass.

#### Blood pressure

Systolic Blood Pressure (SBP) and Diastolic Blood Pressure (DBP) were measured with an aneroid sphygmomanometer (Erka Vario Desk Model), using a stethoscope placed over the brachial artery pulse, proximal and medial to the cubital fossa, and below the bottom edge of the cuff (i.e., 2 cm above the cubital fossa), while the child was seated ^58,59^. Three measurements were taken, and the mean of the 2 last measurements was considered. Then, age-, sex- and height-specific z-scores and percentiles of SBP and DBP were calculated following the recommendations of the American Academy of Pediatrics, and high blood pressure was defined when mean SBP and/or DBP ≥ 90^th^ percentile^58^.

#### Metabolic outcomes

Waist circumference measurements were taken to the nearest 0.1 cm, at the umbilicus level, with the child in a standing position, with the abdomen relaxed, arms at the sides, and feet positioned together. After an overnight fast, a venous blood sample was collected before 11 am. Glucose was measured using ultraviolet (UV) enzymatic assay (hexokinase method), insulin was measured using electrochemiluminescence immunoassay, and triglycerides and high-density lipoprotein cholesterol (HDL cholesterol) were measured using an enzymatic colorimetric assay. Homeostatic model assessment-insulin resistance (HOMA-IR) was calculated as glucose (mg/dL) x insulin (mU/mL)/405 ^60^. Age- and sex-specific percentiles for each parameter were obtained according to the IDEFICS Study ^18^. Metabolic syndrome (MetS) was defined as having ≥ 3 of the following criteria: excess adiposity (waist circumference ≥ 90^th^ percentile), high blood pressure (SBP and/or DBP ≥ 90^th^ percentile), dyslipidemia (triglycerides ≥ 90^th^ or HDL ≤ 10^th^ percentile) and hyperglycemia (HOMA-IR ≥ 90^th^ percentile or fasting glucose ≥ 90^th^ percentile)^18^. Additionally, a continuous metabolic syndrome score combining the components of the metabolic syndrome was calculated, in which higher score levels indicate a worse metabolic profile ^18^.

#### Covariates

Covariates related to both the urban natural spaces exposures and cardiometabolic outcomes were defined according to the literature and depicted in a directed acyclic graph (Figure S2). Maternal educational level reported at birth was measured as the number of completed years of formal schooling and classified according to the International Standard Classification of Education 2011 into low (0-9 years), medium (10-12 years), and high education (>12 years) ^61^. Neighbourhood socioeconomic deprivation index was categorized into quintiles, corresponding the first quintile to the least deprived and the fifth quintile to the most deprived.^62^ Nitrogen dioxide (NO_2_) and particulate matter less than 2.5 μm (PM2.5) were calculated using land use regression (LUR) modelling approach, developed in the European Study of Cohorts for Air Pollution Effects (ESCAPE) framework 2009, and temporal adjustment was conducted using background routine monitoring stations. Neighbourhood socioeconomic deprivation index, NO_2_ and PM2.5 were available at birth, 4, 7 and 10 years of age.

### Statistical analysis

A descriptive analysis of the study population and a comparative analysis between participants and non-participants were performed. The correlation between green and blue spaces exposures, for each and across time-points, was examined and visually displayed using a heat map.

We used latent class mixed models (R package lcmm) to develop trajectories of urban natural spaces exposure from birth until 10 years of age, using a sample with complete data on green and blue space exposure at all time points (n=7549) (Figure S1) ^63^. The Bayesian Information Criteria (BIC) and the Akaike Information Criteria (AIC) were used to identify the optimal number of trajectory groups, and then the mean posterior probability of participants belonging to each latent class was analysed ^64^. We modelled the association of surrounding greenness (NDVI 100m, 250m, and 500m) and distance to the nearest urban green and blue spaces, at each time point and as longitudinal trajectories, with BMI, SBP and DBP z-scores and metabolic syndrome score, continuously, using linear regression models (beta coefficients (β) along with their 95% confidence intervals (CI)), and with overweight/obesity, high blood pressure and metabolic syndrome, as binary outcomes, at 10 years, using logistic regression models (odds ratios (OR), 95% CI). Additionally, we assessed the associations of the urban natural spaces exposure, at each time point and as longitudinal trajectories, with the fat mass index and android-to-gynoid fat ratio using linear regression models. For the analyses with fat mass index, and due to its skewed distribution, we observed a violation of the normality of the residuals assumption in the linear regression models, and thus we log-transformed fat mass index and presented the exponential β and their 95% CI. For each regression analysis, we developed crude and adjusted models including the aforementioned covariates. In the analyses looking at the associations of exposures per time point, we adjusted for neighbourhood socioeconomic deprivation index, NO_2_ and PM2.5 at each respective time point but in the analyses looking at the associations of the trajectories of exposure, due to the high correlation of these environmental data over childhood, we only used information at birth and 10 years. To facilitate the interpretation of effect estimates and to enable comparisons in effect size, we standardized [observed value/standard deviation] the continuous environmental exposures and continuous covariates (PM2.5 and NO_2_). We tested for statistical interactions with child’s sex, maternal education and neighbourhood deprivation index in all models, but none of the interaction terms were statistically significant (p-values > 0.05). We performed sensitivity analyses focused on each of the individual components of the metabolic syndrome, namely glucose, HOMA-IR, triglycerides, HDL, and waist circumference as continuous z-scores. Due to the high number of tests performed, we corrected for multiple testing using the False Discovery Rate (FDR) method ^65^. All analyses were performed in R software version 4.2.0.

## Results

### Study participants

Description of exposure to green and blue spaces from birth until 10 years of age is presented in Tables 1 and S1. The levels of surrounding greenness and distance to the nearest green and blue space showed a small increase from birth until 10 years of age. The mean (standard deviation, SD) NDVI within a buffer of 100m, 250m and 500m ranged from 0.16 (0.08) at birth to 0.21 (0.06) at 10 years old, from 0.20 (0.07) to 0.24 (0.06), and from 0.22 (0.07) to 0.25 (0.06), respectively. The median (25^th^-75^th^ percentile) distance to the nearest green or blue space, from birth until 10 years, ranged from 9.24 (4.94-15.07) hm to 9.53 (5.15-15.19) hm, and from 18.43 (10.44-29.99) hm to 18.85 (10.78-30.39) hm, respectively (Table 1). At each time point, correlations between residential distance to the nearest blue space with residential distance to the nearest green space and surrounding greenness were very low (<0.05) and low (0.10-0.30), respectively (Figure S3). Low to moderate correlations were found between the distance to the nearest green space and surrounding greenness (0.25-0.45). Considering the green and blue space exposures at different time points, moderate to strong correlations were found (0.60-0.95).

**Table 1.** Description of green and blue spaces exposures over childhood (n=4669)

|  | **At birth** | **4 years** | **7 years** | **10 years** |
| --- | --- | --- | --- | --- |
| **NDVI 100m**, mean ± SD | 0.16 ± 0.08 | 0.18 ± 0.07 | 0.21 ± 0.06 | 0.21 ± 0.06 |
| **NDVI 250m**, mean ± SD | 0.20 ± 0.07 | 0.22 ± 0.07 | 0.23 ± 0.06 | 0.24 ± 0.06 |
| **NDVI 500m**, mean ± SD | 0.22 ± 0.07 | 0.24 ± 0.07 | 0.24 ± 0.06 | 0.25 ± 0.06 |
| **Distance to nearest green space (hm)**, median (p25, p75) | 9.24 (4.94, 15.07) | 9.47 (5.06, 15.07) | 9.49 (5.10, 15.14) | 9.53 (5.15, 15.19) |
| **Distance to nearest blue space (hm)**, median (p25, p75) | 18.43 (10.44, 29.99) | 18.65 (10.68, 30.39) | 18.73 (10.69, 30.50) | 18.85 (10.78, 30.39) |

Abbreviations: NDVI, normalized difference vegetation index; SD, standard deviation; hm, hectometre; p25-p75, 25<sup>th</sup> and 75<sup>th</sup> percentiles

Participants’ characteristics regarding covariates and outcomes are shown in Table 2. Females comprise 48.9% of the sample. At 10 years, the prevalence of childhood overweight/obesity, high blood pressure, and metabolic syndrome was 43.4%, 26.5%, and 17.5%, respectively.

**Table 2.** Participants’ characteristics.

|  | **Total (n=4669)** |
| --- | --- |
| **At birth** |  |
| Sex, n (%) |  |
| Female | 2283 (48.9) |
| Male | 2386 (51.1) |
| Maternal educational level, n (%) |  |
| Low | 1988 (42.6) |
| Medium | 1321 (28.3) |
| High | 1360 (29.1) |
| NO<sub>2</sub>, mean ± SD | 38.8 ± 8.4 |
| PM2.5, median (p25, p75) | 21.8 (21.1, 22.5) |
| Neighbourhood socioeconomic deprivation index, n (%) |  |
| 1<sup>st</sup> quintile | 643 (13.8) |
| 2<sup>nd</sup> quintile | 1005 (21.5) |
| 3<sup>rd</sup> quintile | 1186 (25.4) |
| 4<sup>th</sup> quintile | 1029 (22.0) |
| 5<sup>th</sup> quintile | 806 (17.3) |
| **At 4 years** |  |
| NO<sub>2</sub>, mean ± SD | 30.2 ± 6.6 |
| PM2.5, median (p25, p75) | 14.5 (14.0, 15.0) |
| Neighbourhood socioeconomic deprivation index, n (%) |  |
| 1<sup>st</sup> quintile | 658 (14.1) |
| 2<sup>nd</sup> quintile | 1029 (22.0) |
| 3<sup>rd</sup> quintile | 1170 (25.1) |
| 4<sup>th</sup> quintile | 1042 (22.3) |
| 5<sup>th</sup> quintile | 770 (16.5) |
| **At 7 years** |  |
| NO<sub>2</sub>, mean ± SD | 27.3 ± 6.0 |
| PM2.5, median (p25, p75) | 13.8 (13.2, 14.4) |
| Neighbourhood socioeconomic deprivation index, n (%) |  |
| 1<sup>st</sup> quintile | 649 (13.9) |
| 2<sup>nd</sup> quintile | 1054 (22.6) |
| 3<sup>rd</sup> quintile | 1164 (24.9) |
| 4<sup>th</sup> quintile | 1046 (22.4) |
| 5<sup>th</sup> quintile | 756 (16.2) |
| **At 10 years** |  |
| BMI (kg/m<sup>2</sup>), median (p25, p75) | 18.2 (16.3, 20.9) |
| BMI z-scores, median (p25, p75) | 0.7 (-0.2, 1.7) |
| Body weight status |  |
| Normal weight | 2616 (56.6) |
| Overweight/obesity | 2002 (43.4) |
| Fat mass index (kg/m<sup>4</sup>), median (p25, p75) | 3.5 (2.7, 4.8) |
| Android-to-gynoid fat ratio, median (p25, p75) | 0.8 (0.7, 0.9) |
| Systolic BP (mm/Hg), median (p25, p75) | 108.0 (102.0, 114.0) |
| Systolic BP z-score, median (p25, p75) | 0.5 (-0.0, 1.1) |
| Diastolic BP (mm/Hg), median (p25, p75) | 69.0 (64.0, 74.0) |
| Diastolic BP z-score, median (p25, p75) | 0.7 (0.3, 1.1) |
| Blood Pressure |  |
| Normal blood pressure | 3424 (73.5) |
| High blood pressure | 1234 (26.5) |
| Metabolic outcomes |  |
| Waist circumference (cm), median (p25, p75) | 66.0 (60.5, 74.2) |
| Waist circumference z-score, median (p25, p75) | 1.4 (0.4, 2.4) |
| Glucose (mg/dl), median (p25, p75) | 87.0 (83.0, 91.0) |
| Glucose z-score, median (p25, p75) | -0.3 (-0.8, 0.2) |
| Insulin (mU/L), median (p25, p75) | 8.0 (5.5, 11.7) |
| Insulin z-score, median (p25, p75) | 0.6 (-0.3, 1.5) |
| Triglycerides (mg/dl), median (p25, p75) | 60.0 (47.0, 79.0) |
| Triglycerides z-score, median (p25, p75) | 0.6 (0.2, 1.1) |
| HDL cholesterol (mg/dl), median (p25, p75) | 54.0 (48.0, 62.0) |
| HDL cholesterol z-score, median (p25, p75) | -0.1 (-0.6, 0.5) |
| HOMA-IR, median (p25, p75) | 1.7 (1.1, 2.6) |
| HOMA-IR z-score, median (p25, p75) | 0.5 (-0.4, 1.4) |
| Metabolic Syndrome Score, median (p25, p75) | 2.6 (0.7, 4.7) |
| Metabolic Syndrome, n (%) |  |
| No | 2252 (82.5) |
| Yes | 479 (17.5) |
| NO<sub>2</sub>, mean ± SD | 28.2 ± 6.1 |
| PM2.5, median (p25, p75) | 14.1 (13.7, 14.5) |
| Neighbourhood socioeconomic deprivation index, n (%) |  |
| 1<sup>st</sup> quintile | 660 (14.1) |
| 2<sup>nd</sup> quintile | 1042 (22.3) |
| 3<sup>rd</sup> quintile | 1159 (24.8) |
| 4<sup>th</sup> quintile | 1031 (22.1) |
| 5<sup>th</sup> quintile | 777 (16.6) |

Abbreviations: BMI, body mass index; kg/m<sup>2</sup>, kilogram per square metre; SD, standard deviation; p25-p75, 25<sup>th</sup> and 75<sup>th</sup> percentiles; NO<sub>2</sub>, nitrogen dioxide; PM2.5, particulate matter less than 2.5 μm; BP, blood pressure; mmHg, millimetres of mercury; cm, centimetre; mg/dl, milligram per decilitre; mU/L, milliunit per litre

As compared to non-participants, participants had a greater residential distance to the nearest blue space, exhibited a higher exposure to NO_2_ and PM2.5, were less likely to live in a more deprived neighbourhood and their mothers had a higher educational level (Table S2).

### Urban natural spaces per time point and cardiometabolic outcomes

Table 3 shows the associations between green and blue spaces, at each time point, and the cardiometabolic health outcomes at 10 years. We observed an inverse association between the residential distance to the nearest blue space at birth and systolic blood pressure z-scores at 10 years [β (95%CI) = −0.02 (−0.05; −0.00) systolic blood pressure z-score per each standard deviation of distance]. Additionally, a positive association between the residential distance to the nearest green space at 7 and 10 years with metabolic syndrome score at 10 years was found [β (95%CI) = 0.13 (0.01;0.26) and 0.14 (0.01;0.27) metabolic syndrome score per each standard deviation of distance, respectively]. However, after correction for multiple testing, all these associations lost statistical significance. We did not find any statistically significant association for BMI z-score, diastolic blood pressure z-score, overweight/obesity, high blood pressure or metabolic syndrome at 10 years, and for the surrounding greenness at birth, 4, 7 and 10 years with any of the outcomes. The crude associations are depicted in the supplementary material (Table S3). No statistically significant associations were observed between green and blue spaces, at each time point, and the fat mass index and android-to-gynoid fat ratio (Table S4).

**Table 3.**
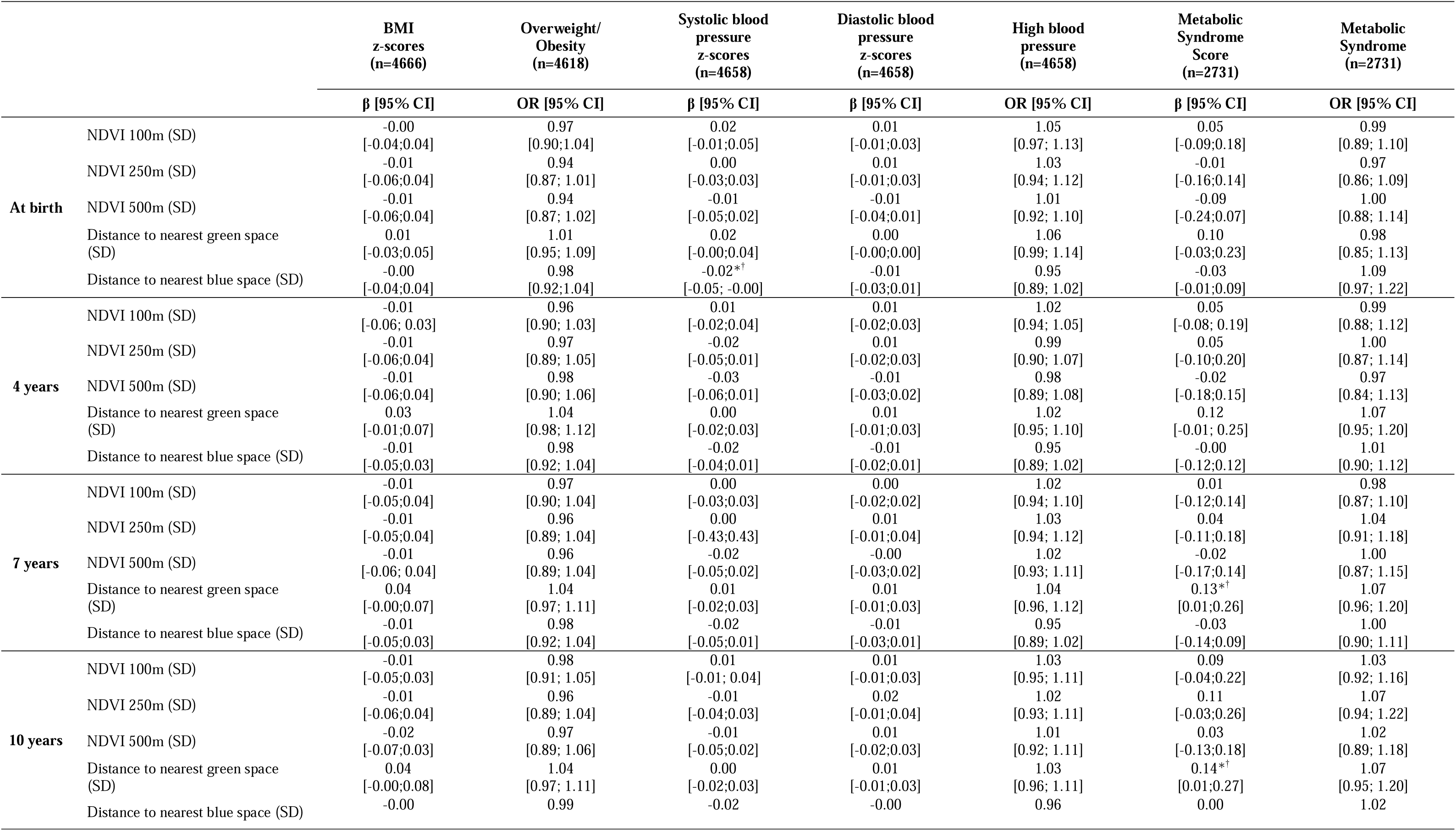
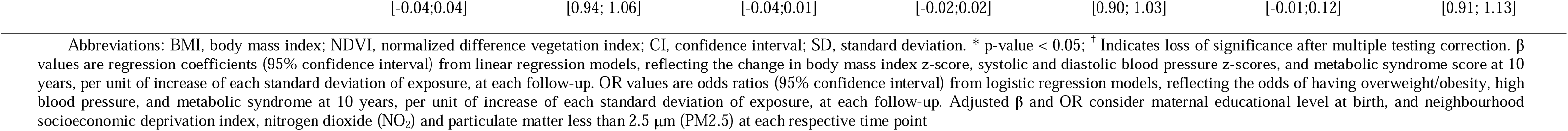
Adjusted linear and logistic regression associations of green and blue spaces, at each time point, with continuous and dichotomic cardiometabolic outcomes at 10 years.

|  |  | **BMI**<br>**z-scores**<br>**(n=4666)** | **Overweight/**<br>**Obesity**<br>**(n=4618)** | **Systolic blood**<br>**pressure**<br>**z-scores**<br>**(n=4658)** | **Diastolic blood**<br>**pressure**<br>**z-scores**<br>**(n=4658)** | **High blood**<br>**pressure**<br>**(n=4658)** | **Metabolic**<br>**Syndrome**<br>**Score**<br>**(n=2731)** | **Metabolic**<br>**Syndrome**<br>**(n=2731)** |
| --- | --- | --- | --- | --- | --- | --- | --- | --- |
|  |  | **β [95% CI]** | **OR [95% CI]** | **β [95% CI]** | **β [95% CI]** | **OR [95% CI]** | **β [95% CI]** | **OR [95% CI]** |
| **At birth** | NDVI 100m (SD) | -0.00<br>[-0.04;0.04] | 0.97<br>[0.90;1.04] | 0.02<br>[-0.01;0.05] | 0.01<br>[-0.01;0.03] | 1.05<br>[0.97; 1.13] | 0.05<br>[-0.09;0.18] | 0.99<br>[0.89; 1.10] |
|  | NDVI 250m (SD) | -0.01<br>[-0.06;0.04] | 0.94<br>[0.87; 1.01] | 0.00<br>[-0.03;0.03] | 0.01<br>[-0.01;0.03] | 1.03<br>[0.94; 1.12] | -0.01<br>[-0.16;0.14] | 0.97<br>[0.86; 1.09] |
|  | NDVI 500m (SD) | -0.01<br>[-0.06;0.04] | 0.94<br>[0.87; 1.02] | -0.01<br>[-0.05;0.02] | -0.01<br>[-0.04;0.01] | 1.01<br>[0.92; 1.10] | -0.09<br>[-0.24;0.07] | 1.00<br>[0.88; 1.14] |
|  | Distance to nearest green space (SD) | 0.01<br>[-0.03;0.05] | 1.01<br>[0.95; 1.09] | 0.02<br>[-0.00;0.04] | 0.00<br>[-0.00;0.00] | 1.06<br>[0.99; 1.14] | 0.10<br>[-0.03;0.23] | 0.98<br>[0.85; 1.13] |
|  | Distance to nearest blue space (SD) | -0.00<br>[-0.04;0.04] | 0.98<br>[0.92;1.04] | -0.02*†<br>[-0.05; -0.00] | -0.01<br>[-0.03;0.01] | 0.95<br>[0.89; 1.02] | -0.03<br>[-0.01;0.09] | 1.09<br>[0.97; 1.22] |
| **4 years** | NDVI 100m (SD) | -0.01<br>[-0.06; 0.03] | 0.96<br>[0.90; 1.03] | 0.01<br>[-0.02;0.04] | 0.01<br>[-0.02;0.03] | 1.02<br>[0.94; 1.05] | 0.05<br>[-0.08; 0.19] | 0.99<br>[0.88; 1.12] |
|  | NDVI 250m (SD) | -0.01<br>[-0.06;0.04] | 0.97<br>[0.89; 1.05] | -0.02<br>[-0.05;0.01] | 0.01<br>[-0.02;0.03] | 0.99<br>[0.90; 1.07] | 0.05<br>[-0.10;0.20] | 1.00<br>[0.87; 1.14] |
|  | NDVI 500m (SD) | -0.01<br>[-0.06;0.04] | 0.98<br>[0.90; 1.06] | -0.03<br>[-0.06;0.01] | -0.01<br>[-0.03;0.02] | 0.98<br>[0.89; 1.08] | -0.02<br>[-0.18;0.15] | 0.97<br>[0.84; 1.13] |
|  | Distance to nearest green space (SD) | 0.03<br>[-0.01;0.07] | 1.04<br>[0.98; 1.12] | 0.00<br>[-0.02;0.03] | 0.01<br>[-0.01;0.03] | 1.02<br>[0.95; 1.10] | 0.12<br>[-0.01; 0.25] | 1.07<br>[0.95; 1.20] |
|  | Distance to nearest blue space (SD) | -0.01<br>[-0.05;0.03] | 0.98<br>[0.92; 1.04] | -0.02<br>[-0.04;0.01] | -0.01<br>[-0.02;0.01] | 0.95<br>[0.89; 1.02] | -0.00<br>[-0.12;0.12] | 1.01<br>[0.90; 1.12] |
| **7 years** | NDVI 100m (SD) | -0.01<br>[-0.05;0.04] | 0.97<br>[0.90; 1.04] | 0.00<br>[-0.03;0.03] | 0.00<br>[-0.02;0.02] | 1.02<br>[0.94; 1.10] | 0.01<br>[-0.12;0.14] | 0.98<br>[0.87; 1.10] |
|  | NDVI 250m (SD) | -0.01<br>[-0.05;0.04] | 0.96<br>[0.89; 1.04] | 0.00<br>[-0.43;0.43] | 0.01<br>[-0.01;0.04] | 1.03<br>[0.94; 1.12] | 0.04<br>[-0.11;0.18] | 1.04<br>[0.91; 1.18] |
|  | NDVI 500m (SD) | -0.01<br>[-0.06; 0.04] | 0.96<br>[0.89; 1.04] | -0.02<br>[-0.05;0.02] | -0.00<br>[-0.03;0.02] | 1.02<br>[0.93; 1.11] | -0.02<br>[-0.17;0.14] | 1.00<br>[0.87; 1.15] |
|  | Distance to nearest green space (SD) | 0.04<br>[-0.00;0.07] | 1.04<br>[0.97; 1.11] | 0.01<br>[-0.02;0.03] | 0.01<br>[-0.01;0.03] | 1.04<br>[0.96; 1.12] | 0.13*†<br>[0.01;0.26] | 1.07<br>[0.96; 1.20] |
|  | Distance to nearest blue space (SD) | -0.01<br>[-0.05;0.03] | 0.98<br>[0.92; 1.04] | -0.02<br>[-0.05;0.01] | -0.01<br>[-0.03;0.01] | 0.95<br>[0.89; 1.02] | -0.03<br>[-0.14;0.09] | 1.00<br>[0.90; 1.11] |
| **10 years** | NDVI 100m (SD) | -0.01<br>[-0.05;0.03] | 0.98<br>[0.91; 1.05] | 0.01<br>[-0.01; 0.04] | 0.01<br>[-0.01;0.03] | 1.03<br>[0.95; 1.11] | 0.09<br>[-0.04;0.22] | 1.03<br>[0.92; 1.16] |
|  | NDVI 250m (SD) | -0.01<br>[-0.06;0.04] | 0.96<br>[0.89; 1.04] | -0.01<br>[-0.04;0.03] | 0.02<br>[-0.01;0.04] | 1.02<br>[0.93; 1.11] | 0.11<br>[-0.03;0.26] | 1.07<br>[0.94; 1.22] |
|  | NDVI 500m (SD) | -0.02<br>[-0.07;0.03] | 0.97<br>[0.89; 1.06] | -0.01<br>[-0.05;0.02] | 0.01<br>[-0.02;0.03] | 1.01<br>[0.92; 1.11] | 0.03<br>[-0.13;0.18] | 1.02<br>[0.89; 1.18] |
|  | Distance to nearest green space (SD) | 0.04<br>[-0.00;0.08] | 1.04<br>[0.97; 1.11] | 0.00<br>[-0.02;0.03] | 0.01<br>[-0.01;0.03] | 1.03<br>[0.96; 1.11] | 0.14*†<br>[0.01;0.27] | 1.07<br>[0.95; 1.20] |
|  | Distance to nearest blue space (SD) | -0.00 | 0.99 | -0.02 | -0.00 | 0.96 | 0.00 | 1.02 |

Abbreviations: BMI, body mass index; NDVI, normalized difference vegetation index; CI, confidence interval; SD, standard deviation. \* p-value < 0.05; <sup>†</sup> Indicates loss of significance after multiple testing correction. $\beta$ values are regression coefficients (95% confidence interval) from linear regression models, reflecting the change in body mass index z-score, systolic and diastolic blood pressure z-scores, and metabolic syndrome score at 10 years, per unit of increase of each standard deviation of exposure, at each follow-up. OR values are odds ratios (95% confidence interval) from logistic regression models, reflecting the odds of having overweight/obesity, high blood pressure, and metabolic syndrome at 10 years, per unit of increase of each standard deviation of exposure, at each follow-up. Adjusted $\beta$ and OR consider maternal educational level at birth, and neighbourhood socioeconomic deprivation index, nitrogen dioxide (NO<sub>2</sub>) and particulate matter less than 2.5 μm (PM<sub>2.5</sub>) at each respective time point

### Urban natural spaces exposure trajectories

The values of AIC and BIC used to identify the optimal number of trajectory groups, and the mean posterior probability of participants belonging to each latent class are depicted in Tables S5 and S6, respectively. We identified the following distinct trajectories from birth to 10 years: A) for NDVI within 100m: 1) high stable (n=747) and 2) low ascending (n=3922); B) for NDVI within 250m: 1) high stable (n=1091), 2) low ascending (n=3223), 3) ascending (n=207), 4) descending (n=148); C) for NDVI within 500m: 1) high stable (n=1287), 2) low ascending (n=3007), 3) ascending (n=227), 4) descending (n=148); D) for the distance to the nearest green space: 1) medium stable (n=4396), 2) ascending (n=137), 3) descending (n=136); and E) for the distance to the nearest blue space: 1) high stable (n=4141), 2) low stable (n=528) (Figure 1, Table S7).

**Figure 1.**
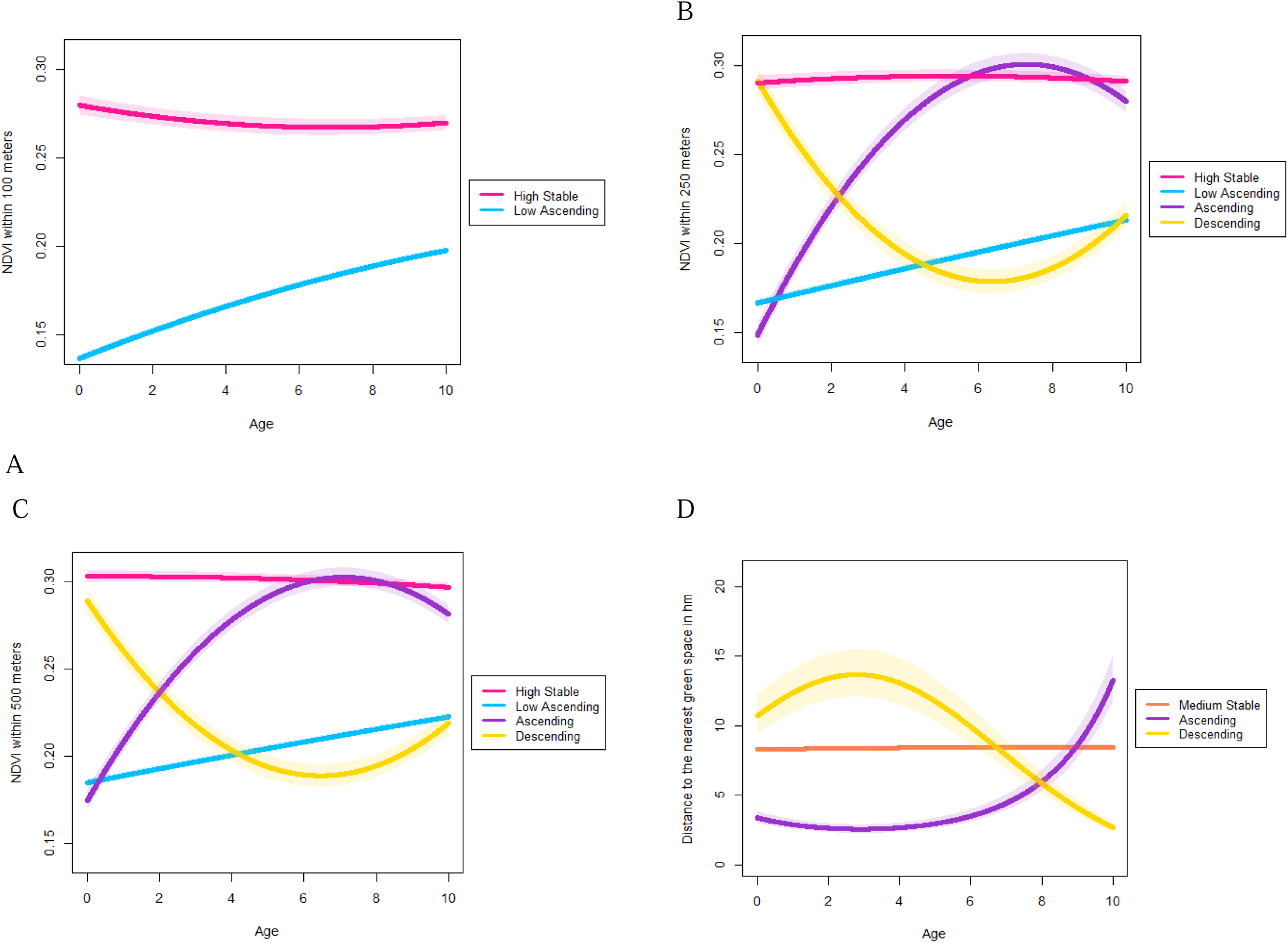
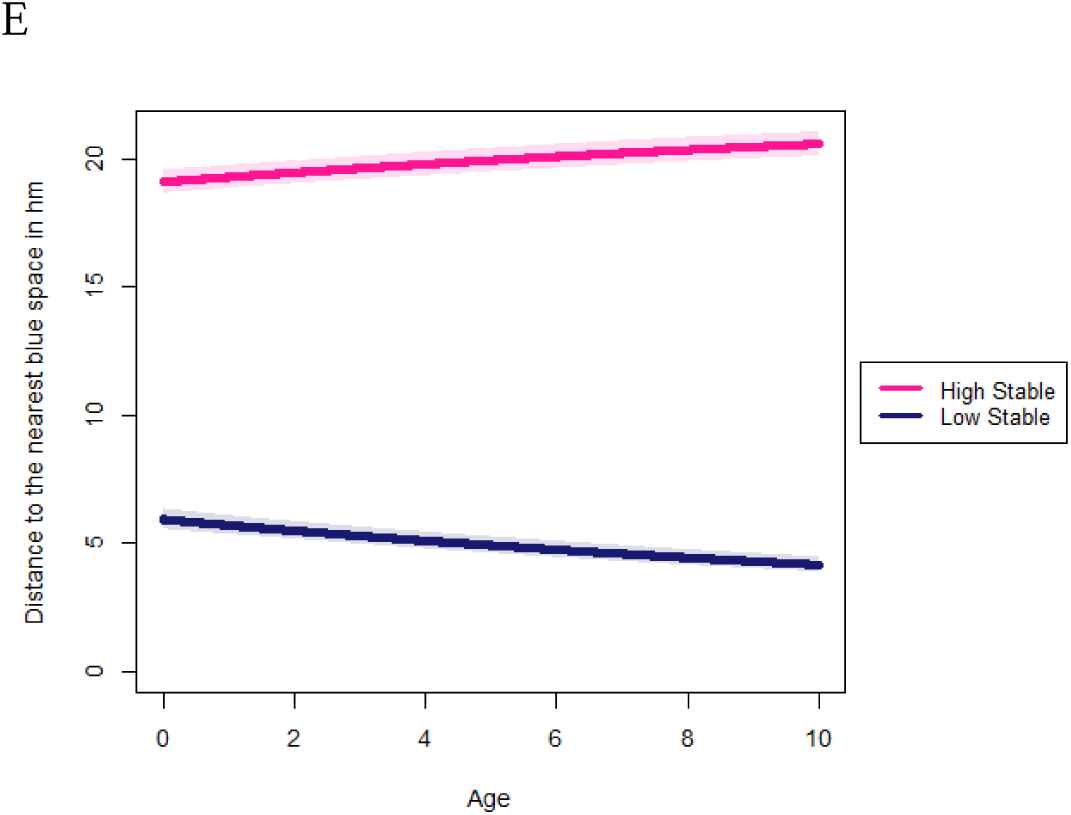
NDVI 100m, 250m and 500m, and residential distance to green and blue spaces, from birth to 10 years of age. (A) Normalized Difference Vegetation Index (NDVI) trajectories within 100m of the child residence, (B) NDVI trajectories within 250m of the child residence, (C) NDVI trajectories within 500m of the child residence, (D) Residential distance to the nearest green space (hm) trajectories, (E) Residential distance to the nearest blue space (hm) trajectories from birth to 10 years

### Urban natural spaces exposure trajectories and cardiometabolic outcomes

Table 4 shows the associations between green and blue spaces exposure trajectories and the cardiometabolic health outcomes at 10 years. We observed that, as compared to children in the high stable trajectory of NDVI within 500m, those in the descending trajectory of NDVI within 500m presented a lower diastolic blood pressure z-score at 10 years [β (95%CI) = −0.15 (−0.26; −0.04)] as well as a lower metabolic syndrome score at 10 years [β (95%CI) = −0.72 (−1.38; −0.07)]. Nonetheless, after correction for multiple testing, these associations lost statistical significance. We did not find any statistically significant association for any other cardiometabolic outcome at 10 years and any other green and blue spaces exposure trajectory. The crude associations are depicted in the supplementary material (Table S8). No statistically significant associations were observed between green and blue spaces exposure trajectories and the fat mass index and android-to-gynoid fat ratio (Table S9).

**Table 4.**
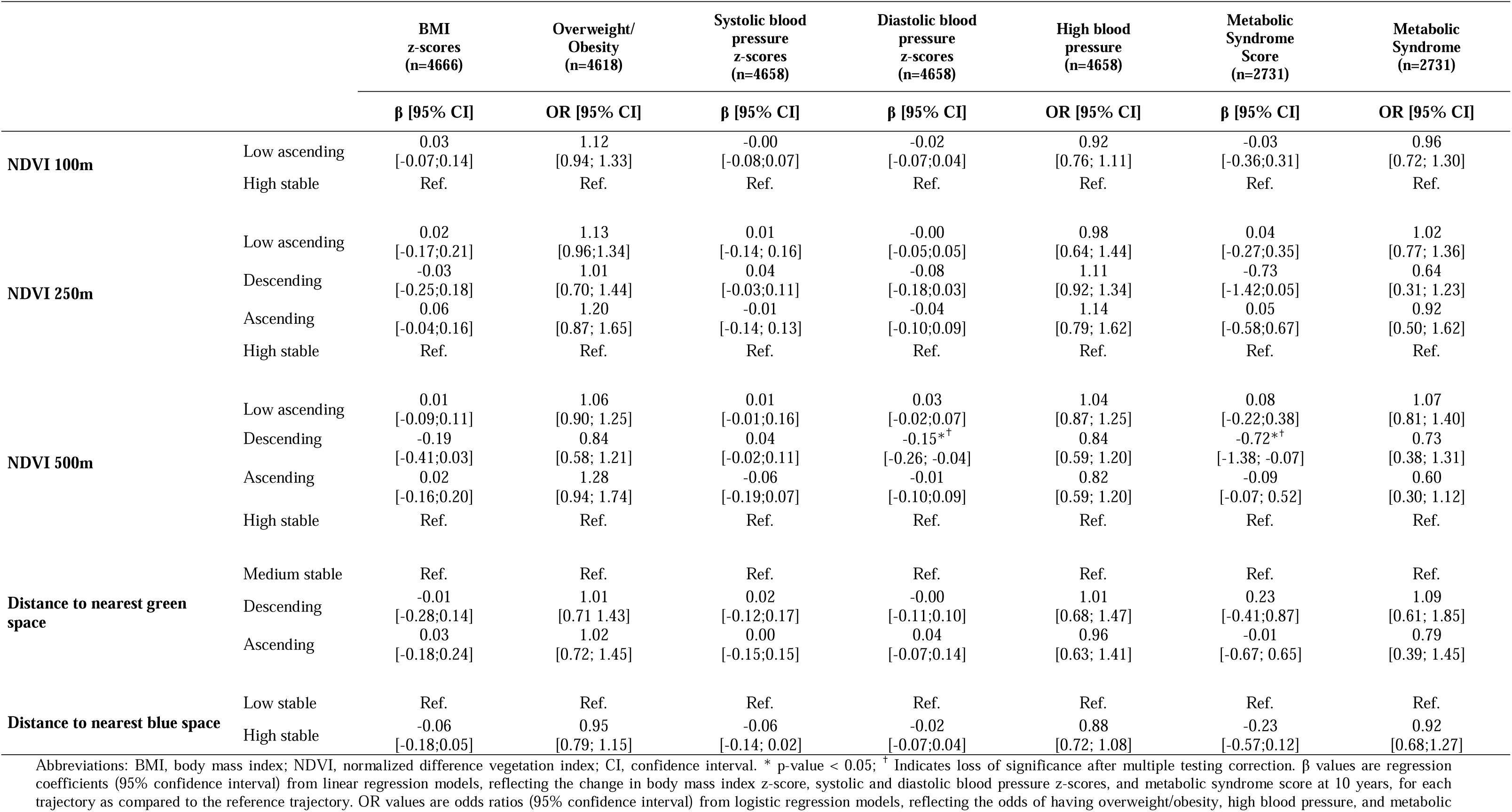
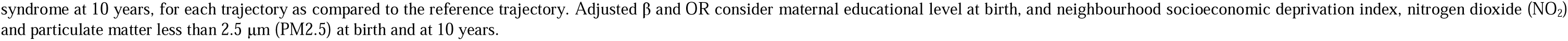
Adjusted linear and logistic regression associations of green and blue spaces trajectories with continuous and dichotomic cardiometabolic outcomes at 10 years.

### Sensitivity analysis

Tables S10 and S11 show the associations of exposure to green and blue spaces, at each time point and as longitudinal trajectories, respectively, with each of the individual components of the metabolic syndrome at 10 years as continuous z-scores. We observed that higher distance to the nearest blue space at birth, 4, 7, and 10 years was associated with lower glucose z-scores while higher distance to the nearest green space at 4 years was associated with higher HOMA-IR z-scores (p-values<0.05). We also observed that higher distance to the nearest blue space at 7 years was associated with lower triglycerides z-scores and higher surrounding greenness within 250m at 10 years was associated with lower HDL cholesterol z-scores (p-values<0.05). Additionally, we observed that, as compared to children in the low stable trajectory of distance to the nearest blue space, those in the high stable trajectory presented lower glucose z-scores and, as compared to children in the high stable trajectory of NDVI within 500m, those in the descending trajectory presented lower waist circumference z-scores (p-values<0.05). All these associations lost statistical significance after correction for multiple testing.

## Discussion

This study did not support our hypothesis that a greater exposure to green and blue spaces is associated with a better cardiometabolic health in childhood. We did not observe significant associations of urban green and blue spaces with body mass index, body fat content and fat distribution at 10 years. The observed associations for blood pressure and metabolic outcomes were of small magnitude, not present after correction for multiple testing, and were not consistently observed across time points and between trajectories for continuous and dichotomic outcomes.

Two previous longitudinal studies with large sample sizes - 51 873 children and adolescents aged 6 to 19 years old in the United States of America and 79 992 children followed-up until 5 years old in Spain - have shown associations between increased neighbourhood greenness (NDVI) and decreased BMI z-scores^37,39^. Likewise, a cross-sectional study using data of the 2018 Health Behaviour in School-aged Children (HBSC) Italian survey, among a regionally representative sample of 2065 Italian adolescents aged 11 to 13 years, found that higher exposure to greenness (NDVI) was associated with a lower risk of obesity^39^. Two cross-sectional studies among 1,489 Lithuaniás children and 442 Turkish children have found no significant association between the distance to the nearest green space and the risk of overweight or obesity ^26,27^, while a cross-sectional study including 440 primary school children aged 6-13 years old in Kathmandu metropolitan city (Nepal) found a higher risk of overweight and obesity among children whose residence was > 1km from the nearest green space^36^. In an individual-participant data (IPD) meta-analysis of over 35,000 children from 10 European birth cohorts across eight countries, residential green space exposure using NDVI within a 300 m buffer and proximity to green spaces during prenatal and childhood periods were not associated with BMI z-scores^42^. Further studies that defined green space by caregiver-reported access to parks and recreational spaces^29^ or by time spent in green spaces^28^ found no association with body mass index. On the other hand, a Norwegian cross-sectional study with 10,527 participants aged between 14 and 16 years found that the odds of having overweight were 1.38 times higher for those living in the greenest areas compared to those in the least green areas^40^. Studies looking at the exposure to blue spaces in relation to body mass index are scarce. Baseline data from the Whole of Systems Trial of Prevention Strategies for Childhood Obesity (WHOSTOPS), with data from 1216 children aged 8.5-13 years, found that female children living closer to the coastal area had lower odds of having overweight/obesity ^19,20^. In the present study, we found no significant association of surrounding greenness and residential distance to the nearest green or blue space, at any time point and when using trajectories from birth to 10 years with BMI z-score, overweight/obesity and body fat content and distribution.

Cross-sectional studies from Austria^32^, Italy^32^, China^31,33^ and Germany^34^, from 1251 to 588,004 children and adolescents samples, showed that higher greenness around school^31,32^ and residence^34^ was associated with lower systolic ^31,32,34^ and diastolic blood pressure ^31,34^, reduced odds of hypertension ^31^, whereas reduced greenness was associated with higher blood pressure ^34^. A Chinese longitudinal and dynamic cohort of children and adolescents (2005-2018) found that a decrease in greenness around schools was associated with higher blood pressure^33^. Yet, other studies did not find such associations. The HELIX study, a consortium of six birth cohorts, comprising 4279 children aged 4 to 5 years old found that neither the exposure to NDVI (100m, 300m, 500m) nor the distance to the nearest major green and blue space during pregnancy, were significantly related with both systolic and diastolic blood pressure in middle-childhood^43^. Similarly, the Childhood and Adolescence Surveillance and Prevention of Adult Non-communicable disease study [CASPIA-IV] (2015) – which included 12 340 children aged 7 to 18 years old, living in urban and rural areas of Iran, found no association between subjective proximity to green spaces (perception of having access to such spaces within a 15-minute walk from their home) and systolic and diastolic blood pressure ^66^. The previously mentioned IPD meta-analysis also did not find an association between residential green space exposure and systolic and diastolic blood pressure^42^. In the present study, after correction for multiple testing, no associations were observed between surrounding greenness and residential distance to the nearest green or blue space, at any time point and when using trajectories from birth to 10 years, with blood pressure z-scores and high blood pressure.

To date, no study was conducted to evaluate the association of urban green and blue spaces on metabolic syndrome in the paediatric population. The metabolic syndrome is a cluster of conditions that occur together, namely excess body fat around the waist, high blood pressure, dyslipidemia and hyperglycemia. In the present study, after correction for multiple testing, no significant association was observed between the exposure to blue and green spaces and metabolic syndrome or its components.

Cardiometabolic conditions are complex and result from a web of risk versus protective factors. Our findings suggest that urban natural spaces may not play a role in its development. Green and blue spaces have been associated with salutogenic effects, particularly related with its utility on the development of health-promoting behaviours. These spaces might allow greater opportunities for recreational physical activity and walkability, social contacts and community engagement, reduction of stressors exposure and cognitive and psychological restoration^4^. Despite the theoretical framework, the effect of urban natural spaces by itself may not be sufficient to exert an impact on cardiometabolic health in children. However, we cannot disregard some methodological particularities of our study that might have precluded us from finding significant associations. We considered blue spaces as only larger water bodies such as rivers and seas, thereby overlooking the potential positive impacts of smaller water bodies like fountains, ponds, and lakes ^67^, and we had information on greenness and residential proximity to the nearest green spaces rather than utilization, which might be influenced by additional factors such as accessibility, infrastructure, and safety ^68^. It has been shown that the impact of green spaces on health differ across socioeconomic groups, and that they are differently distributed according to neighbourhood socioeconomic characteristics ^69,70^. For instance, green spaces are frequently lacking in the most economically disadvantaged neighbourhoods ^69,71^. A study conducted in Porto municipality revealed that in socially deprived neighbourhoods, green spaces often have more safety issues, signs of damage, insufficient recreational equipment, as well as fewer amenities like seating, restrooms, and cafes ^69^, potentially making them less appealing to children and their families. In this study, there was no interaction with maternal education and neighbourhood deprivation index, but we cannot disregard that these variables, used as a proxy for socioeconomic position, may have been insufficient to depict the nature of potential differences in socioeconomic characteristics over time ^72^. Also, this study focuses on residential urban green and blue space exposures and thus, it might have been affected by the Uncertain Geographic Context Problem, since focusing on residential neighbourhoods does not account for the time that individuals spend outside their home environment ^73^. For instance, we did not account for the fact that school-aged children, primarily from primary education onwards, spend at least eight hours per day at school or engaged in related activities from Monday to Friday. Therefore, the urban green and blue spaces surrounding school environments may play a more significant role. Indeed, an association between accessibility to green spaces near schools and health improvements was observed in seven-year-old children from G21^74^. One could argue that the choice of multiple testing correction was overly conservative, which may explain why no significant associations were found. Indeed, for metabolic outcomes there were associations prior to the correction for multiple testing, yet they were of low magnitude and lacked consistency across time points and between trajectories.

This study has strengths worth acknowledging. The use of data from a large population-based birth cohort study enabled the systematic collection of data throughout time. Additionally, the prevalence of overweight/obesity, high blood pressure and metabolic syndrome in our sample was higher than the prevalence reported in childhood worldwide ^15–17^, assuring a robust sample to explore the tested associations. The combination of both blue and green spaces along with two different measures to assess green space – greenness (NDVI) and residential distance to the nearest green space - offer a more comprehensive approach for analysing these features within the built environment. For greenness, we used different buffer sizes of NDVI to capture various dimensions of exposure. For instance, a larger buffer size (500m) might include commuting spaces and spaces where children spend their time^75^ while smaller buffer sizes (250m and 100m) may reflect the exposure immediately surrounding children’s residences^12^ Also, the availability of these data from birth until late childhood allowed us to employ a longitudinal approach, assessing their association with cardiometabolic outcomes per follow-up and as life-course trajectories, as opposed and adding to most previous evidence from cross-sectional studies. Moreover, we used detailed cardiometabolic health outcomes, which were defined using robust data coming from anthropometric and blood pressure measurements as well as from imaging techniques and fasting blood samples conducted by trained healthcare professionals following standardized procedures.

However, this study has some limitations. We lacked data to assess the use of green and blue spaces by children enrolled in G21 cohort. In our study, the exclusion of participants with missing data on both exposure and outcome measures may have introduced selection bias. When comparing excluded versus included participants, we observed that among the excluded 58% had mothers with a lower educational level (vs 42.6% included) and 25% lived in most deprived areas classified in the 5th quintile of the neighbourhood deprivation index (vs 17%). Thus, our sample might not truly represent the socioeconomic characteristics of the initial cohort participants. Moreover, we did not have data on other potential confounders such as noise pollution, which could also impact our results through residual confounding.

In conclusion, this study did not find robust associations between urban green and blue spaces over key developmental periods and body mass index, body fat content and distribution, blood pressure and metabolic outcomes in childhood, and thus does not support the hypothesized beneficial impact of these natural spaces on cardiometabolic health.

## Acknowledgments

We gratefully acknowledge the families enrolled in Generation XXI for their kindness, the participating hospitals and their staff for their help and support, and all previous and current members of the research and field team for their enthusiasm and perseverance.

## Financial Support

Generation XXI (G21) was funded by Programa Operacional de Saúde – Saúde XXI, Quadro Comunitário de Apoio III and Administração Regional de Saúde Norte (Regional Department of Ministry of Health). This work was supported by FCT - Fundação para a Ciência e Tecnologia, I.P. through the projects with references UIDB/04750/2020 and LA/P/0064/2020 and DOI identifiers https://doi.org/10.54499/UIDB/04750/2020 and https://doi.org/10.54499/LA/P/0064/2020. Environmental exposures were assessed within the EXALAR 21 project (PTDC/GES-AMB/30193/2017) funded by the European Regional Development Fund (FEDER), through the Competitiveness and Internationalization Operational Programme, and by national funding from the Foundation for Science and Technology (FCT). Ana Isabel Ribeiro was supported by National Funds through FCT, under the programme of ‘Stimulus of Scientific Employment – Individual Support’ within the contract CEECIND/02386/2018 (https://doi.org/10.54499/CEECIND/02386/2018/CP1538/CT0001). Susana Santos was supported by the European Uniońs Horizon Europe Research and Innovation Programme under the Marie Sklodowska-Curie Postdoctoral Fellowship Grant Agreement No. 101109136 (URBANE). Views and opinions expressed are however those of the authors only and do not necessarily reflect those of the European Union or the European Research Executive Agency (REA). Neither the European Union nor the granting authority can be held responsible for them. This study was supported by the European Union Horizon 2020 Research and Innovation Programme under Grant Agreement 824989 (EUCAN-Connect) and 874583 (ATHLETE).

## Author contributions

BV was responsible for data analysis and interpretation, and for drafting the manuscript. BA was responsible for data analysis and interpretation. RP was responsible for drafting the manuscript and revision. AIR was responsible for data acquisition and curation of environmental data, critical revision as well as funding acquisition for EXALAR 21 project. HB was responsible for critical revision of the manuscript for intellectual content. SS was responsible for study concept and design, acquisition of data and critical revision of the manuscript for intellectual content and final approval of the version to be published. All authors approved the final manuscript as submitted and agreed to be accountable for all aspects of the work.

## Conflict of interest

The authors declare no competing interests.

## Data Availability Statement

The data from Generation XXI are not publicly available due to privacy or ethical restrictions. The data can be made available for research proposals on request to the Generation XXI Executive Committee. Further information about Generation XXI can be obtained via the Generation XXI website [www.geracao21.com] or by emailing.

## Ethical Standards disclosure

All phases of the study complied with the Ethical Principles for Medical Research Involving Human Subjects expressed in the Declaration of Helsinki. The baseline and follow-up evaluations until 10 years were approved by the University of Porto Medical School/ S. João Hospital Centre Ethics Committee. At baseline and follow-up evaluations, all procedures were explained to participants, and informed consent was signed by one of the parents or legal guardians. The baseline evaluation was additionally approved by the Data Protection National Commission and the study follows the present EU General Data Protection Regulation under close supervision of the Data Protection Office of Institute of Public Health of the University of Porto (ISPUP).

## Notes

### Competing Interest Statement

The authors have declared no competing interest.

### Funding Statement

Generation XXI (G21) was funded by Programa Operacional de Saude: Saude XXI, Quadro Comunitario de Apoio III and Administracao Regional de Saude Norte (Regional Department of Ministry of Health). This work was supported by FCT - Fundacao para a Ciencia e Tecnologia, I.P. through the projects with references UIDB/04750/2020 and LA/P/0064/2020 and DOI identifiers https://doi.org/10.54499/UIDB/04750/2020 and https://doi.org/10.54499/LA/P/0064/2020. Environmental exposures were assessed within the EXALAR 21 project (PTDC/GES-AMB/30193/2017) funded by the European Regional Development Fund (FEDER), through the Competitiveness and Internationalization Operational Programme, and by national funding from the Foundation for Science and Technology (FCT). Ana Isabel Ribeiro was supported by National Funds through FCT, under the programme of Stimulus of Scientific Employment: Individual Support within the contract CEECIND/02386/2018 (https://doi.org/10.54499/CEECIND/02386/2018/CP1538/CT0001). Susana Santos was supported by the European Union Horizon Europe Research and Innovation Programme under the Marie Sklodowska-Curie Postdoctoral Fellowship Grant Agreement No. 101109136 (URBANE). This study was supported by the European Union Horizon 2020 Research and Innovation Programme under Grant Agreement 824989 (EUCAN-Connect) and 874583 (ATHLETE).

### Author Declarations

All phases of the study complied with the Ethical Principles for Medical Research Involving Human Subjects expressed in the Declaration of Helsinki. The baseline and follow-up evaluations until 10 years were approved by the University of Porto Medical School/ S. Joao Hospital Centre Ethics Committee. At baseline and follow-up evaluations, all procedures were explained to participants, and informed consent was signed by one of the parents or legal guardians. The baseline evaluation was additionally approved by the Data Protection National Commission and the study follows the present EU General Data Protection Regulation under close supervision of the Data Protection Office of Institute of Public Health of the University of Porto (ISPUP).

## References

1. World Health Organization. Urban Health. https://www.who.int/news-room/fact-sheets/detail/urban-health

2. World Health Organization. Integrating health in urban and territorial planning: a sourcebook. 2020.

3. Smith N, Georgiou M, King AC, Tieges Z, Webb S, Chastin S. Urban blue spaces and human health: A systematic review and meta-analysis of quantitative studies. Cities. 2021/12/01/ 2021;119:103413. 10.1016/j.cities.2021.103413

4. Nieuwenhuijsen MJ, Khreis H, Triguero-Mas M, Gascon M, Dadvand P. Fifty Shades of Green: Pathway to Healthy Urban Living. Epidemiology. Jan 2017;28(1):63–71. doi:10.1097/ede.0000000000000549

5. Van den Bosch M, Sang ÅO. Urban natural environments as nature-based solutions for improved public health–A systematic review of reviews. Environmental research. 2017;158:373–384.

6. World Health Organization t. Global recommendations on physical activity for health. World Health Organization; 2010.

7. Markevych I, Schoierer J, Hartig T, et al. Exploring pathways linking greenspace to health: Theoretical and methodological guidance. Environmental research. 2017;158:301–317.

8. White MP, Elliott LR, Gascon M, Roberts B, Fleming LE. Blue space, health and well-being: A narrative overview and synthesis of potential benefits. Environmental research. 2020;191:110169.

9. Steptoe A, Kivimäki M. Stress and cardiovascular disease. Nature Reviews Cardiology. 2012;9(6):360–370.

10. Kubzansky LD, Huffman JC, Boehm JK, et al. Positive psychological well-being and cardiovascular disease: JACC health promotion series. Journal of the American College of Cardiology. 2018;72(12):1382–1396.

11. Gunawardena KR, Wells MJ, Kershaw T. Utilising green and bluespace to mitigate urban heat island intensity. Science of the Total Environment. 2017;584:1040–1055.

12. James P, Banay RF, Hart JE, Laden F. A Review of the Health Benefits of Greenness. Curr Epidemiol Rep. Jun 2015;2(2):131–142. doi:10.1007/s40471-015-0043-7

13. UNICEF. The State of the World’s Children 2012: Children in an urban world. 2012.

14. Goryakin Y, Rocco L, Suhrcke M. The contribution of urbanization to non-communicable diseases: Evidence from 173 countries from 1980 to 2008. Econ Hum Biol. Aug 2017;26:151–163. doi:10.1016/j.ehb.2017.03.004

15. World Health Organization. Obesity and overweight - fact sheet 2018 https://www.who.int/news-room/fact-sheets/detail/obesity-and-overweight.

16. Song P, Zhang Y, Yu J, et al. Global Prevalence of Hypertension in Children: A Systematic Review and Meta-analysis. JAMA Pediatr. Dec 1 2019;173(12):1154–1163. doi:10.1001/jamapediatrics.2019.3310

17. Weihe P, Weihrauch-Blüher S. Metabolic Syndrome in Children and Adolescents: Diagnostic Criteria, Therapeutic Options and Perspectives. Curr Obes Rep. Dec 2019;8(4):472–479. doi:10.1007/s13679-019-00357-x

18. Ahrens W, Moreno LA, Mårild S, et al. Metabolic syndrome in young children: definitions and results of the IDEFICS study. Int J Obes (Lond). Sep 2014;38 Suppl 2:S4–14. doi:10.1038/ijo.2014.130

19. Crooks N, Becker D, Gaskin CJ, et al. Relationship between “Blue Space” Proximity and Children’s Weight Status, Health Behaviors, and Health-Related Quality of Life among a Sample of Regional Victorian Primary School Children. Child Obes. Oct 2022;18(7):494–506. doi:10.1089/chi.2021.0219

20. Wood SL, Demougin PR, Higgins S, Husk K, Wheeler BW, White M. Exploring the relationship between childhood obesity and proximity to the coast: A rural/urban perspective. Health Place. Jul 2016;40:129–36. doi:10.1016/j.healthplace.2016.05.010

21. Sanders T, Feng X, Fahey PP, Lonsdale C, Astell-Burt T. Greener neighbourhoods, slimmer children? Evidence from 4423 participants aged 6 to 13 years in the Longitudinal Study of Australian children. Int J Obes (Lond). Aug 2015;39(8):1224–9. doi:10.1038/ijo.2015.69

22. Dadvand P, Villanueva CM, Font-Ribera L, et al. Risks and benefits of green spaces for children: a cross-sectional study of associations with sedentary behavior, obesity, asthma, and allergy. Environ Health Perspect. Dec 2014;122(12):1329–35. doi:10.1289/ehp.1308038

23. Lovasi GS, Schwartz-Soicher O, Quinn JW, et al. Neighborhood safety and green space as predictors of obesity among preschool children from low-income families in New York City. Prev Med. Sep 2013;57(3):189–93. doi:10.1016/j.ypmed.2013.05.012

24. Yang Y, Jiang Y, Xu Y, Mzayek F, Levy M. A cross-sectional study of the influence of neighborhood environment on childhood overweight and obesity: Variation by age, gender, and environment characteristics. Prev Med. Mar 2018;108:23–28. doi:10.1016/j.ypmed.2017.12.021

25. Liu GC, Wilson JS, Qi R, Ying J. Green neighborhoods, food retail and childhood overweight: differences by population density. Am J Health Promot. Mar-Apr 2007;21(4 Suppl):317–25. doi:10.4278/0890-1171-21.4s.317

26. Petraviciene I, Grazuleviciene R, Andrusaityte S, Dedele A, Nieuwenhuijsen MJ. Impact of the Social and Natural Environment on Preschool-Age Children Weight. Int J Environ Res Public Health. Mar 5 2018;15(3) doi:10.3390/ijerph15030449

27. Akpinar A. Urban green spaces for children: A cross-sectional study of associations with distance, physical activity, screen time, general health, and overweight. Urban Forestry & Urban Greening. 2017/07/01/ 2017;25:66–73. 10.1016/j.ufug.2017.05.006

28. Benjamin-Neelon SE, Platt A, Bacardi-Gascon M, Armstrong S, Neelon B, Jimenez-Cruz A. Greenspace, physical activity, and BMI in children from two cities in northern Mexico. Prev Med Rep. Jun 2019;14:100870. doi:10.1016/j.pmedr.2019.100870

29. Alexander DS, Huber LR, Piper CR, Tanner AE. The association between recreational parks, facilities and childhood obesity: a cross-sectional study of the 2007 National Survey of Children’s Health. J Epidemiol Community Health. May 2013;67(5):427–31. doi:10.1136/jech-2012-201301

30. Xiao X, Yang BY, Hu LW, et al. Greenness around schools associated with lower risk of hypertension among children: Findings from the Seven Northeastern Cities Study in China. Environ Pollut. Jan 2020;256:113422. doi:10.1016/j.envpol.2019.113422

31. Luo Y-N, Yang B-Y, Zou Z, et al. Associations of greenness surrounding schools with blood pressure and hypertension: A nationwide cross-sectional study of 61,229 children and adolescents in China. Environmental Research. 2022/03/01/ 2022;204:112004. 10.1016/j.envres.2021.112004

32. Dzhambov AM, Lercher P, Markevych I, Browning M, Rüdisser J. Natural and built environments and blood pressure of Alpine schoolchildren. Environ Res. Mar 2022;204(Pt A):111925. doi:10.1016/j.envres.2021.111925

33. Chen L, Xie J, Ma T, et al. Greenness alleviates the effects of ambient particulate matter on the risks of high blood pressure in children and adolescents. Sci Total Environ. Mar 15 2022;812:152431. doi:10.1016/j.scitotenv.2021.152431

34. Markevych I, Thiering E, Fuertes E, et al. A cross-sectional analysis of the effects of residential greenness on blood pressure in 10-year old children: results from the GINIplus and LISAplus studies. BMC Public Health. May 20 2014;14:477. doi:10.1186/1471-2458-14-477

35. Warembourg C, Nieuwenhuijsen M, Ballester F, et al. Urban environment during early-life and blood pressure in young children. Environ Int. Jan 2021;146:106174. doi:10.1016/j.envint.2020.106174

36. Manandhar S, Suksaroj TT, Rattanapan C. The Association between Green Space and the Prevalence of Overweight/Obesity among Primary School Children. Int J Occup Environ Med. Jan 2019;10(1):1–10. doi:10.15171/ijoem.2019.1425

37. Daniels KM, Schinasi LH, Auchincloss AH, Forrest CB, Roux AVD. The built and social neighborhood environment and child obesity: A systematic review of longitudinal studies. Preventive Medicine. 2021;153:106790.

38. Bellisario V, Comoretto RI, Berchialla P, et al. The Association between Greenness and Urbanization Level with Weight Status among Adolescents: New Evidence from the HBSC 2018 Italian Survey. Int J Environ Res Public Health. May 12 2022;19(10) doi:10.3390/ijerph19105897

39. de Bont J, Hughes R, Tilling K, et al. Early life exposure to air pollution, green spaces and built environment, and body mass index growth trajectories during the first 5 years of life: A large longitudinal study. Environ Pollut. Nov 2020;266(Pt 3):115266. doi:10.1016/j.envpol.2020.115266

40. Wilhelmsen CK, Skalleberg K, Raanaas RK, Tveite H, Aamodt G. Associations between green area in school neighbourhoods and overweight and obesity among Norwegian adolescents. Prev Med Rep. Sep 2017;7:99–105. doi:10.1016/j.pmedr.2017.05.020

41. Potwarka LR, Kaczynski AT, Flack AL. Places to play: association of park space and facilities with healthy weight status among children. Journal of community health. 2008;33:344–350.

42. Fernandes A, Avraam D, Cadman T, et al. Green spaces and respiratory, cardiometabolic, and neurodevelopmental outcomes: An individual-participant data meta-analysis of >35.000 European children. Environ Int. Jun 28 2024;190:108853. doi:10.1016/j.envint.2024.108853

43. Warembourg C, Nieuwenhuijsen M, Ballester F, et al. Urban environment during early-life and blood pressure in young children. Environment International. 2021;146:106174.

44. Loomis D, Kromhout H. Exposure variability: concepts and applications in occupational epidemiology. Am J Ind Med. Jan 2004;45(1):113–22. doi:10.1002/ajim.10324

45. Halfon N, Hochstein M. Life course health development: an integrated framework for developing health, policy, and research. Milbank Q. 2002;80(3):433–79, iii. doi:10.1111/1468-0009.00019

46. Larsen PS, Kamper-Jørgensen M, Adamson A, et al. Pregnancy and birth cohort resources in europe: a large opportunity for aetiological child health research. Paediatr Perinat Epidemiol. Jul 2013;27(4):393–414. doi:10.1111/ppe.12060

47. Alves E, Correia S, Barros H, Azevedo A. Prevalence of self-reported cardiovascular risk factors in Portuguese women: a survey after delivery. International journal of public health. 2012;57:837–847.

48. World Medical Association. Declaration of Helsinki. 2013. http://www.wma.net/en/20activities/10ethics/10helsinki/

49. Instituto Nacional de Estatística (INE). População residente por local de residência (resultados preliminares Censos 2021). https://ine.pt/xportal/xmain?xpid=INE&xpgid=ine_indicadores&indOcorrCod=0010745&contexto=pi&selTab=tab0

50. URBACT. Porto Metropolitan Area. https://urbact.eu/articles/porto-metropolitan-area

51. Madureira H, Andresen T, Monteiro A. Green structure and planning evolution in Porto. Urban Forestry & Urban Greening. 2011;10(2):141–149.

52. Paciência I, Moreira A, Moreira C, et al. Neighbourhood green and blue spaces and allergic sensitization in children: A longitudinal study based on repeated measures from the Generation XXI cohort. Science of The Total Environment. 2021/06/10/ 2021;772:145394. 10.1016/j.scitotenv.2021.145394

53. Ribeiro AI, Santos AC, Vieira VM, Barros H. Hotspots of childhood obesity in a large metropolitan area: does neighbourhood social and built environment play a part? Int J Epidemiol. Jun 1 2020;49(3):934–943. doi:10.1093/ije/dyz205

54. Cavaleiro Rufo J, Paciência I, Hoffimann E, Moreira A, Barros H, Ribeiro AI. The neighbourhood natural environment is associated with asthma in children: A birth cohort study. Allergy. Jan 2021;76(1):348–358. doi:10.1111/all.14493

55. de Onis M, Onyango AW, Borghi E, Siyam A, Nishida C, Siekmann J. Development of a WHO growth reference for school-aged children and adolescents. Bull World Health Organ. Sep 2007;85(9):660–7. doi:10.2471/blt.07.043497

56. Santos S, Severo M, Lopes C, Oliveira A. Anthropometric Indices Based on Waist Circumference as Measures of Adiposity in Children. Obesity (Silver Spring). May 2018;26(5):810–813. doi:10.1002/oby.22170

57. Wells JC, Cole TJ. Adjustment of fat-free mass and fat mass for height in children aged 8 y. Int J Obes Relat Metab Disord. Jul 2002;26(7):947–52. doi:10.1038/sj.ijo.0802027

58. National High Blood Pressure Education Program Working Group on High Blood Pressure in Children and Adolescents. The fourth report on the diagnosis, evaluation, and treatment of high blood pressure in children and adolescents. Pediatrics. Aug 2004;114(2 Suppl 4th Report):555–76.

59. Miranda JO, Cerqueira RJ, Barros H, Areias JC. Maternal Diabetes Mellitus as a Risk Factor for High Blood Pressure in Late Childhood. Hypertension. 2019;73(1):e1–e7. doi:doi:10.1161/HYPERTENSIONAHA.118.11761

60. Matthews DR, Hosker JP, Rudenski AS, Naylor BA, Treacher DF, Turner RC. Homeostasis model assessment: insulin resistance and beta-cell function from fasting plasma glucose and insulin concentrations in man. Diabetologia. Jul 1985;28(7):412–9. doi:10.1007/bf00280883

61. UNESCO Institute for Statistics. International standard classification of education: ISCED 2011. Comparative Social Research. 2012;30

62. Ribeiro AI, Launay L, Guillaume E, Launoy G, Barros H. The Portuguese version of the European Deprivation Index: Development and association with all-cause mortality. PLoS One. 2018;13(12):e0208320. doi:10.1371/journal.pone.0208320

63. Proust-Lima C, Philipps V, Liquet B. Estimation of extended mixed models using latent classes and latent processes: the R package lcmm. arXiv preprint arXiv:150300890. 2015;

64. Lennon H, Kelly S, Sperrin M, et al. Framework to construct and interpret latent class trajectory modelling. BMJ open. 2018;8(7):e020683.

65. Glickman ME, Rao SR, Schultz MR. False discovery rate control is a recommended alternative to Bonferroni-type adjustments in health studies. Journal of clinical epidemiology. 2014;67(8):850–857.

66. Abbasi B, Pourmirzaei M, Hariri S, et al. Subjective Proximity to Green Spaces and Blood Pressure in Children and Adolescents: The CASPIAN-V Study. J Environ Public Health. 2020;2020:8886241. doi:10.1155/2020/8886241

67. Georgiou M, Morison G, Smith N, Tieges Z, Chastin S. Mechanisms of Impact of Blue Spaces on Human Health: A Systematic Literature Review and Meta-Analysis. Int J Environ Res Public Health. Mar 3 2021;18(5) doi:10.3390/ijerph18052486

68. Schipperijn J, Bentsen P, Troelsen J, Toftager M, Stigsdotter UK. Associations between physical activity and characteristics of urban green space. Urban Forestry & Urban Greening. 2013/01/01/ 2013;12(1):109–116. 10.1016/j.ufug.2012.12.002

69. Hoffimann E, Barros H, Ribeiro AI. Socioeconomic Inequalities in Green Space Quality and Accessibility-Evidence from a Southern European City. Int J Environ Res Public Health. Aug 15 2017;14(8) doi:10.3390/ijerph14080916

70. Kabisch N. The influence of socio-economic and socio-demographic factors in the association between urban green space and health. Biodiversity and health in the face of climate change. 2019:91–119.

71. Astell-Burt T, Feng X, Mavoa S, Badland HM, Giles-Corti B. Do low-income neighbourhoods have the least green space? A cross-sectional study of Australia’s most populous cities. BMC Public Health. 2014/03/31 2014;14(1):292. doi:10.1186/1471-2458-14-292

72. Almeida DQ, Paciência I, Moreira C, et al. Green and blue spaces and lung function in the Generation XXI cohort: a life-course approach. European Respiratory Journal. 2022;60(6)

73. Park YM, Kwan M-P. Multi-contextual segregation and environmental justice research: Toward fine-scale spatiotemporal approaches. International journal of environmental research and public health. 2017;14(10):1205.

74. Ribeiro AI, Tavares C, Guttentag A, Barros H. Association between neighbourhood green space and biological markers in school-aged children. Findings from the Generation XXI birth cohort. Environment International. 2019/11/01/ 2019;132:105070. 10.1016/j.envint.2019.105070

75. Pearce JR. Complexity and uncertainty in geography of health research: incorporating life-course perspectives. Annals of the American Association of Geographers. 2018;108(6):1491–1498.

