## supplements for "Residential exposure to green and blue spaces over childhood and cardiometabolic health outcomes: The Generation XXI birth cohort"

(2) Laboratório para a Investigação Integrativa e Translacional em Saúde Populacional (ITR), Universidade do Porto, Rua das Taipas, n° 135, 4050-600 Porto, Portugal

(3) Departamento de Saúde Pública e Ciências Forenses e Educação Médica, Faculdade de Medicina, Universidade do Porto, Alameda Prof. Hernâni Monteiro, 4200-319, Porto, Portugal

**Content**

**Figure S1.** Flowchart of the study population

**Figure S2**. Directed acyclic graph (DAG) to identify potential confounders of the association between residential green and blue spaces with child cardiometabolic health

**Figure S3**. Correlation matrix of the environmental exposures

**Table S1.** Description of green and blue space exposures over childhood in the sample used to derive trajectories

**Table S2.** Comparison between included and non-included participants

**Table S3**. Crude linear and logistic regression associations between green and blue spaces, at each time point, with continuous and dichotomic cardiometabolic outcomes at 10 years

**Table S4** Crude and adjusted linear regression associations of green and blue spaces, at each time point, with continuous body fat content and fat distribution outcomes at 10 years

**Table S5.** Criteria to assess model fit for latent group analysis model for NDVI 100m, NDVI 250m and NDVI 500m, and distance to the nearest green and blue spaces.

**Table S6**. Probability of belonging to each latent class for NDVI 100m, NDVI 250m, and NDVI 500m and distance to the nearest green and blue space

**Table S7.** Description of the natural spaces exposure trajectories

**Table S8**. Crude linear and logistic regression associations between green and blue spaces trajectories with continuous and dichotomic cardiometabolic outcomes at 10 years

**Table S9.** Crude and adjusted linear regression associations of green and blue spaces trajectories with continuous body fat content and fat distribution outcomes at 10 years

**­­­­­­Table S10.** Adjusted linear regression associations of green and blue spaces, at each time point, with continuous z-scores of metabolic syndrome components at 10 years

**Table S11**. Adjusted linear regression associations between green and blue spaces trajectories with continuous z-scores of metabolic syndrome components at 10 years

**Figure S1**. Flowchart of the study population

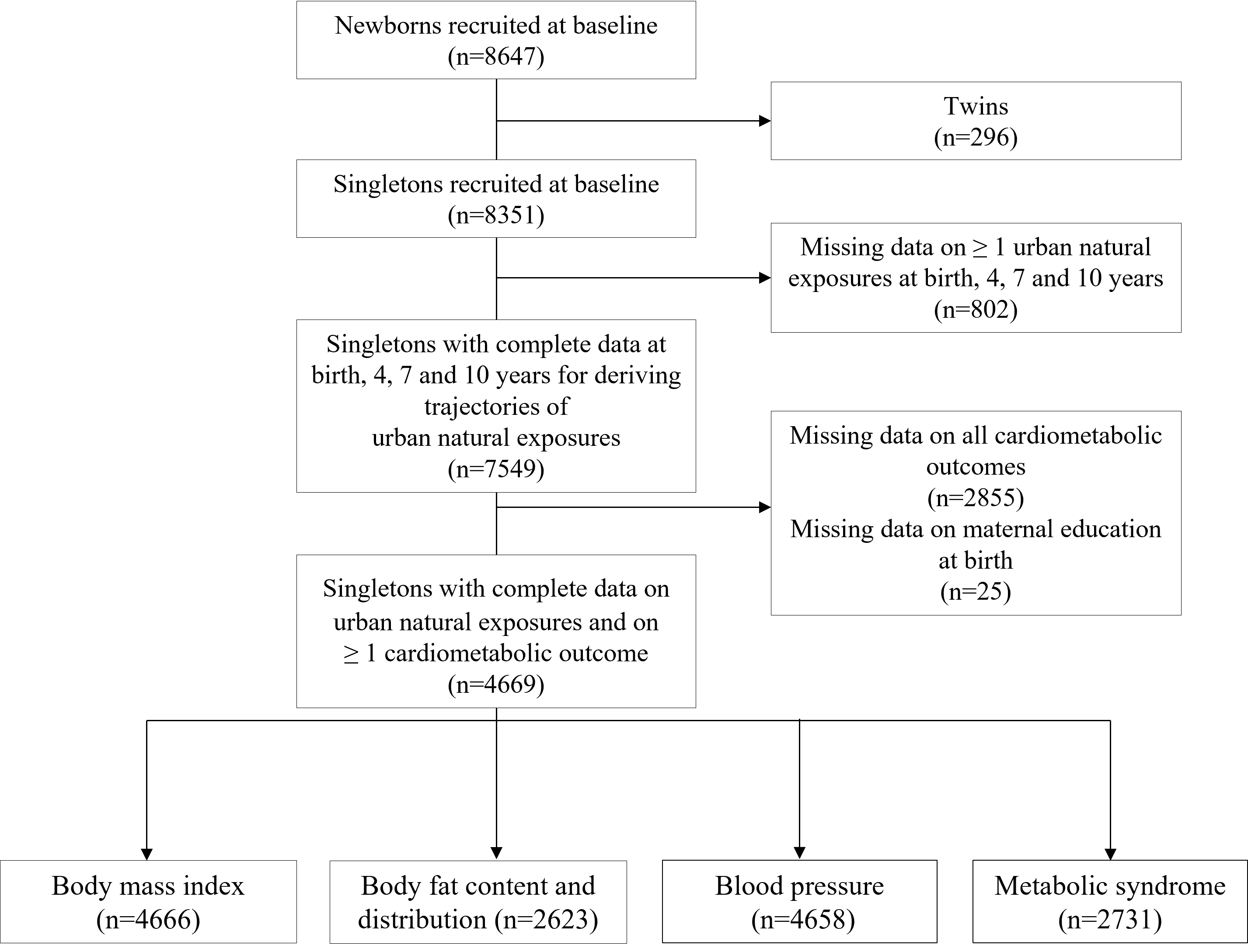

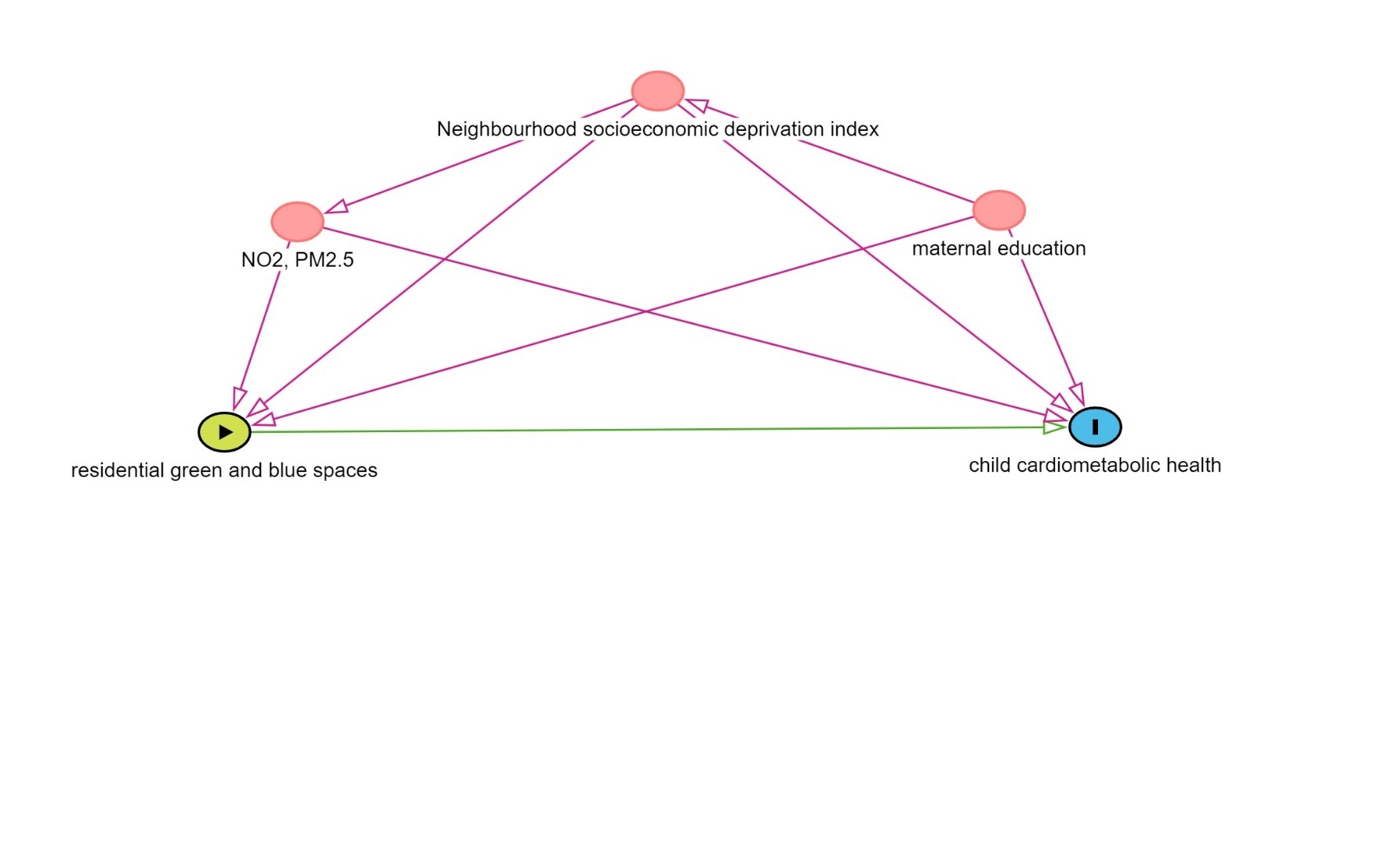
**Figure S2**. Directed acyclic graph (DAG) to identify potential confounders of the association between residential green and blue spaces and child cardiometabolic health

**Figure S3**. Correlation matrix of the environmental exposures

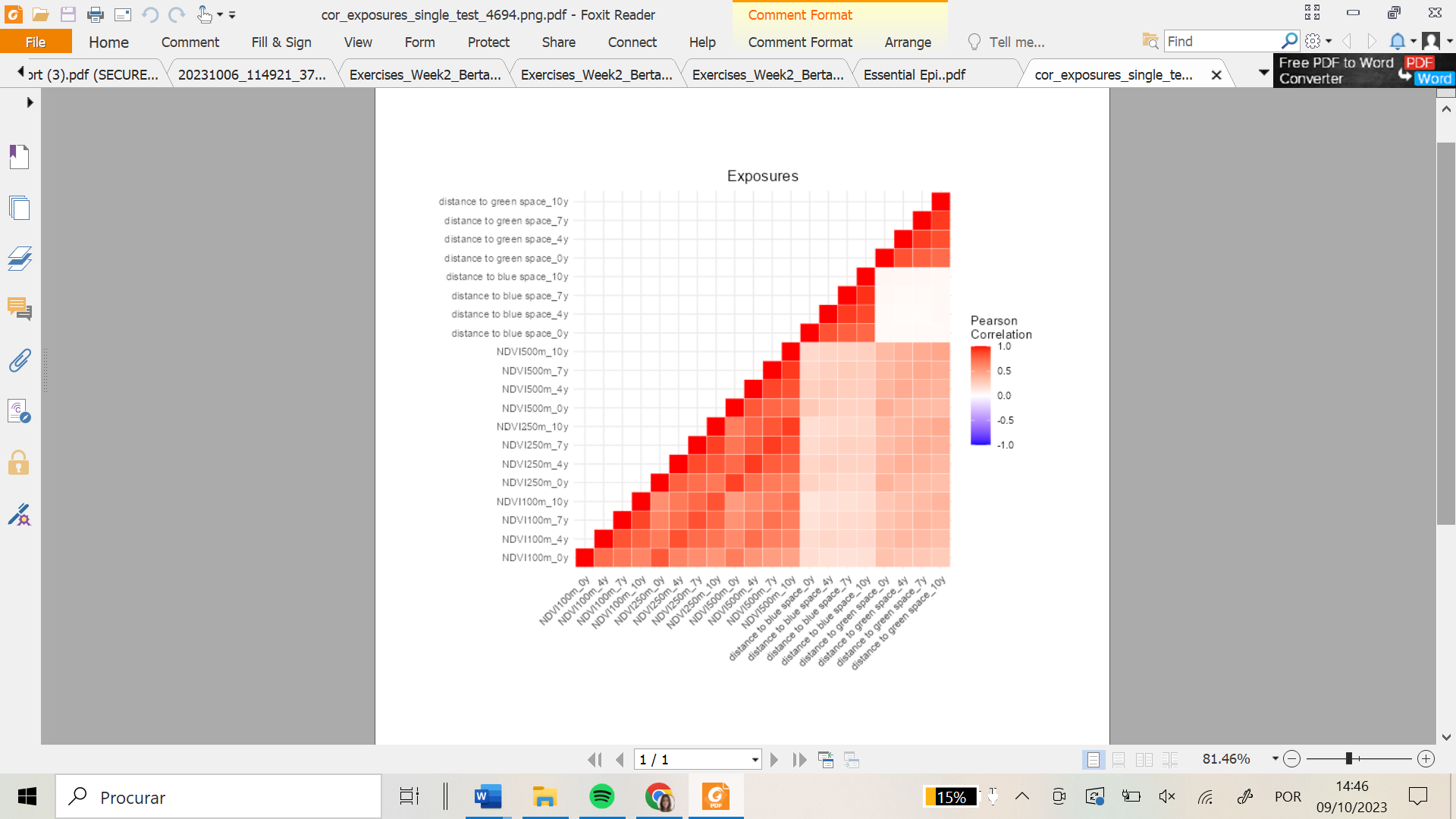

**Table S1**. Description of green and blue space exposures over childhood in the sample used to derive trajectories (n=7549)

|  | **At birth** | **4 years** | **7 years** | **10 years** |
| --- | --- | --- | --- | --- |
| **NDVI 100m**, mean ± SD | 0.16 ± 0.08 | 0.18 ± 0.07 | 0.21 ± 0.06 | 0.21 ± 0.06 |
| **NDVI 250m**, mean ± SD | 0.20 ± 0.08 | 0.21 ± 0.07 | 0.23 ± 0.06 | 0.23 ± 0.06 |
| **NDVI 500m**, mean ± SD | 0.22 ± 0.07 | 0.23 ± 0.07 | 0.24 ± 0.06 | 0.25 ± 0.06 |
| **Distance to nearest green space (hm)**, median (p25, p75) | 9.30  (4.92, 14.90) | 9.54  (5.02, 14.91) | 9.60  (5.11, 14.95) | 9.62  (5.15, 14.98) |
| **Distance to nearest blue space (hm)**, median (p25, p75) | 18.43  (10.27, 29.90) | 18.51  (10.44, 30.24) | 18.57  (10.44, 30.23) | 18.66  (10.49, 30.10) |
| Abbreviations: NDVI, normalized difference vegetation index; SD, standard deviation; p25-p75, 25^th^ and 75^th^ percentiles | | | | |

**Table S2**. Comparison between included and non-included participants

|  | **Included**  (n=4669) | **Non-included**  (n=3682) | p-value |
| --- | --- | --- | --- |
| **Environmental exposures** |  |  |  |
| *At birth* |  |  |  |
| NDVI 100m, mean ± SD | 0.16 ± 0.08 | 0.16 ± 0.08 | 0.529 |
| NDVI 250m, mean ± SD | 0.20 ± 0.07 | 0.20 ± 0.08 | **0.031** |
| NDVI 500m, mean ± SD | 0.22 ± 0.07 | 0.22 ± 0.07 | **0.002** |
| Distance to nearest green space (hm), median (p25, p75) | 9.24  (4.94, 15.07) | 9.09  (4.70, 14.64) | 0.189 |
| Distance to nearest blue space (hm), median (p25, p75) | 18.43  (10.44, 29.99) | 18.00  (9.36, 29.44) | **0.018** |
| *4 years* |  |  |  |
| NDVI 100m, mean ± SD | 0.18 ± 0.07 | 0.18 ± 0.07 | 0.991 |
| NDVI 250m, mean ± SD | 0.22 ± 0.07 | 0.21 ± 0.08 | **0.006** |
| NDVI 500m, mean ± SD | 0.24 ± 0.07 | 0.23 ± 0.07 | **<0.001** |
| Distance to nearest green space (hm), median (p25, p75) | 9.47  (5.06, 15.07) | 9.53  (4.88, 14.68) | 0.326 |
| Distance to nearest blue space (hm), median (p25, p75) | 18.65  (10.68, 30.39) | 17.98  (9.37, 29.37) | **0.005** |
| *7 years* |  |  |  |
| NDVI 100m, mean ± SD | 0.21 ± 0.06 | 0.20 ± 0.06 | **0.010** |
| NDVI 250m, mean ± SD | 0.23 ± 0.06 | 0.23 ± 0.06 | **<0.001** |
| NDVI 500m, mean ± SD | 0.24 ± 0.06 | 0.24 ± 0.06 | **<0.001** |
| Distance to nearest green space (hm), median (p25, p75) | 9.49  (5.10, 15.14) | 9.56  (4.98, 14.62) | 0.396 |
| Distance to nearest blue space (hm), median (p25, p75) | 18.73  (10.69, 30.50) | 17.97  (9.39, 29.19) | **0.002** |
| *10 years* |  |  |  |
| NDVI 100m, mean ± SD | 0.21 ± 0.06 | 0.21 ± 0.06 | **0.038** |
| NDVI 250m, mean ± SD | 0.24 ± 0.06 | 0.23 ± 0.06 | **0.005** |
| NDVI 500m, mean ± SD | 0.25 ± 0.06 | 0.24 ± 0.06 | **<0.001** |
| Distance to nearest green space (hm), median (p25, p75) | 9.53  (5.15, 15.19) | 9.56  (4.99, 14.69) | 0.395 |
| Distance to nearest blue space (hm), median (p25, p75) | 18.85  (10.78, 30.39) | 17.97  (9.37, 29.31) | **0.001** |
| **Participant characteristics** |  |  |  |
| *At birth* |  |  |  |
| Sex |  |  |  |
| Female | 2283 (48.9) | 1803 (49.0) | 0.966 |
| Male | 2386 (51.1) | 1879 (51.0) |  |
| **Maternal characteristics** |  |  |  |
| Maternal educational level, n (%) |  |  |  |
| Low | 1988 (42.6) | 2111 (58.2) | **<0.001** |
| Medium | 1321 (28.3) | 897 (24.7) |  |
| High | 1360 (29.1) | 618 (17.0) |  |
| **Environmental characteristics** |  |  |  |
| *At birth* |  |  |  |
| NO_2_, mean ± SD | 38.8 ± 8.4 | 37.5 ± 9.9 | **<0.001** |
| PM2.5, median (p25, p75) | 21.8  (21.1, 22.5) | 21.6  (20.6, 22.4) | **<0.001** |
| Neighbourhood socioeconomic deprivation index, n (%) |  |  |  |
| 1^st^ quintile | 643 (13.8) | 404 (11.1) | **<0.001** |
| 2^nd^ quintile | 1005 (21.5) | 621 (17.0) |  |
| 3^rd^ quintile | 1186 (25.4) | 871 (23.9) |  |
| 4^th^ quintile | 1029 (22.0) | 829 (22.7) |  |
| 5^th^ quintile | 806 (17.3) | 924 (25.3) |  |
| *4 years* |  |  |  |
| NO_2_, mean ± SD | 30.2 ± 6.6 | 28.8 ± 7.8 | **<0.001** |
| PM2.5, median (p25, p75) | 14.5  (14.0, 15.0) | 14.4  (13.6, 15.0) | **<0.001** |
| Neighbourhood socioeconomic deprivation index, n (%) |  |  |  |
| 1^st^ quintile | 658 (14.1) | 413 (11.3) | **<0.001** |
| 2^nd^ quintile | 1029 (22.0) | 619 (17.0) |  |
| 3^rd^ quintile | 1170 (25.1) | 864 (23.7) |  |
| 4^th^ quintile | 1042 (22.3) | 864 (23.7) |  |
| 5^th^ quintile | 770 (16.5) | 891 (24.4) |  |
| *7 years* |  |  |  |
| NO_2_, mean ± SD | 27.3 ± 6.0 | 26.1 ± 7.1 | **<0.001** |
| PM2.5, median (p25, p75) | 13.8  (13.2, 14.4) | 13.6  (12.8, 14.3) | **<0.001** |
| Neighbourhood socioeconomic deprivation index, n (%) |  |  |  |
| 1^st^ quintile | 649 (13.9) | 422 (11.6) | **<0.001** |
| 2^nd^ quintile | 1054 (22.6) | 627 (17.2) |  |
| 3^rd^ quintile | 1164 (24.9) | 859 (23.5) |  |
| 4^th^ quintile | 1046 (22.4) | 860 (23.6) |  |
| 5^th^ quintile | 756 (16.2) | 881 (24.1) |  |
| *10 years* |  |  |  |
| NO_2_, mean ± SD | 28.2 ± 6.1 | 26.8 ± 7.4 | **<0.001** |
| PM2.5, median (p25, p75) | 14.1  (13.7, 14.5) | 13.9  (13.1, 14.4) | **<0.001** |
| Neighbourhood socioeconomic deprivation index, n (%) |  |  |  |
| 1^st^ quintile | 660 (14.1) | 428 (11.7) | **<0.001** |
| 2^nd^ quintile | 1042 (22.3) | 633 (17.4) |  |
| 3^rd^ quintile | 1159 (24.8) | 871 (23.9) |  |
| 4^th^ quintile | 1031 (22.1) | 844 (23.1) |  |
| 5^th^ quintile | 777 (16.6) | 870 (23.9) |  |
| Abbreviations: NDVI, Normalized Difference Vegetation Index; hm, hectometre; SD, standard deviation; p25, p75, 25^th^ and 75^th^ percentiles; NO_2_, nitrogen dioxide; PM2.5, particulate matter less than 2.5 µm | | | |

**Table S3**. Crude linear and logistic regression associations between green and blue spaces, at each time point, with continuous and dichotomic cardiometabolic outcomes at 10 years

|  | | **BMI**  **z-scores**  **(n=4666)** | **Overweight/**  **Obesity**  **(n=4618)** | **Systolic blood pressure**  **z-scores**  **(n=4658)** | **Diastolic blood pressure**  **z-scores**  **(n=4658)** | **High blood**  **pressure**  **(n=4658)** | **Metabolic**  **Syndrome Score**  **(n=2731)** | **Metabolic**  **Syndrome**  **(n=2731)** |
| --- | --- | --- | --- | --- | --- | --- | --- | --- |
|  |  | **β [CI 95%]** | **OR [CI 95%]** | **β [CI 95%]** | **β [CI 95%]** | **OR [CI 95%]** | **β [CI 95%]** | **OR [CI 95%]** |
| **At birth** | NDVI 100m (SD) | 0.03  [-0.00; 0.07] | 1.02  [0.96; 1.08] | 0.03*  [0.01; 0.06] | 0.02  [-0.00; 0.03] | 1.07  [1.00; 1.14] | 0.12*  [0.00; 0.23] | 1.00  [0.90; 1.08] |
|  | NDVI 250 m (SD) | 0.04*  [0.00; 0.08] | 1.02  [0.96; 1.08] | 0.02  [-0.01; 0.04] | 0.02  [-0.00; 0.03] | 1.05  [0.98; 1.12] | 0.09  [-0.02; 0.20] | 1.03  [0.93; 1.13] |
|  | NDVI 500m (SD) | 0.04*  [0.01; 0.08] | 1.02  [0.96; 1.08] | 0.01  [-0.02; 0.03] | 0.01  [-0.01; 0.02] | 1.03  [0.97; 1.10] | 0.05  [-0.06; 0.16] | 1.01  [0.92; 1.12] |
|  | Distance to nearest green space (SD) | 0.04*  [0.01; 0.08] | 1.05  [0.99; 1.12] | 0.02  [-0.00; 0.05] | 0.02  [0.00; 0.04] | 1.07  [1.00; 1.15] | 0.17^*^  [0.05; 0.29] | 1.10  [1.00; 1.12] |
|  | Distance to nearest blue space (SD) | 0.00  [-0.03; 0.04] | 0.98  [0.93; 1.04] | -0.02  [-0.05; 0.00] | -0.01  [-0.03; 0.01] | 0.95  [0.89; 1.02] | -0.01  [-0.12; 0.11] | 0.99  [0.89; 1.09] |
| **4 years** | NDVI 100m (SD) | 0.02  [-0.01; 0.06] | 1.00  [0.94, 1.06] | 0.02  [-0.00; 0.05] | 0.01  [-0.01; 0.03] | 1.05  [0.98; 1.12] | 0.12*  [0.01; 0.23] | 1.01  [0.91; 1.12] |
|  | NDVI 250 m (SD) | 0.03  [-0.00; 0.07] | 1.02  [0.96; 1.08] | 0.00  [-0.02; 0.03] | 0.02  [-0.00; 0.03] | 1.03  [0.97; 1.10] | 0.11*  [0.00; 0.23] | 1.01  [0.92; 1.12] |
|  | NDVI 500m (SD) | 0.04*  [0.00; 0.07] | 1.03  [0.97; 1.09] | -0.00  [-0.03; 0.03] | 0.01  [-0.01; 0.03] | 1.03  [0.96; 1.10] | 0.08  [-0.03; 0.19] | 1.00  [0.90; 1.10] |
|  | Distance to nearest green space (SD) | 0.05*  [0.02; 0.09] | 1.06  [1.00; 1.13] | 0.02  [-0.01; 0.04] | 0.01  [-0.00; 0.03] | 1.05  [0.98; 1.10] | 0.18  [0.07; 0.30] | 1.08  [0.98; 1.19] |
|  | Distance to nearest blue space (SD) | -0.00  [-0.04; 0.03] | 0.99  [0.93; 1.05] | -0.02  [-0.05; 0.01] | -0.01  [-0.02; 0.01] | 0.95  [0.89; 1.02] | 0.01  [-0.11; 0.12] | 0.99  [0.90; 1.10] |
| **7 years** | NDVI 100m (SD) | 0.02  [-0.01; 0.06] | 1.01  [0.95; 1.07] | 0.02  [-0.01; 0.04] | 0.01  [-0.01; 0.03] | 1.04  [0.98; 1.12] | 0.09  [-0.02; 0.20] | 1.01  [0.91; 1.11] |
|  | NDVI 250 m (SD) | 0.03  [-0.00; 0.07] | 1.01  [0.96; 1.08] | 0.01  [-0.01; 0.04] | 0.02  [-0.00; 0.04] | 1.05  [0.99; 1.12] | 0.12*  [0.00; 0.23] | 1.03  [0.94; 1.14] |
|  | NDVI 500m (SD) | 0.03  [-0.00; 0.07] | 1.02  [0.96; 1.08] | 0.01  [-0.02; 0.03] | 0.01  [-0.01; 0.03] | 1.04  [0.97; 1.11] | 0.08  [-0.03; 0.19] | 1.01  [0.91; 1.11] |
|  | Distance to nearest green space (SD) | 0.06*  [0.02; 0.09] | 1.06  [1.00; 1.12] | 0.02  [-0.01; 0.04] | 0.01  [-0.00; 0.03] | 1.05  [0.99; 1.12] | 0.20*  [0.08; 0.31] | 1.09  [0.98; 1.20] |
|  | Distance to nearest blue space (SD) | -0.00  [-0.04; 0.03] | 0.99  [0.93; 1.05] | -0.02  [-0.04; 0.01] | -0.01  [-0.03; 0.01] | 0.95  [0.89; 1.02] | -0.01  [-0.13; 0.10] | 0.99  [0.89; 1.09] |
| **10 years** | NDVI 100m (SD) | 0.03  [-0.01; 0.06] | 1.01  [0.95; 1.06] | 0.02  [-0.01; 0.04] | 0.01  [-0.01; 0.03] | 1.03  [0.96; 1.09] | 0.12*  [0.00; 0.23] | 1.01  [0.92; 1.12] |
|  | NDVI 250 m (SD) | 0.03  [-0.00; 0.07] | 1.01  [0.96; 1.08] | 0.01  [-0.02; 0.03] | 0.01  [-0.00; 0.03] | 1.02  [0.96; 1.09] | 0.13*  [0.01; 0.24] | 1.03  [0.93; 1.13] |
|  | NDVI 500m (SD) | 0.03  [-0.01; 0.06] | 1.02  [0.96; 1.08] | 0.01  [-0.02; 0.03] | 0.01  [-0.01; 0.03] | 1.02  [0.95; 1.09] | 0.07  [-0.04; 0.18] | 1.00  [0.90; 1.10] |
|  | Distance to nearest green space (SD) | 0.06*  [0.03; 0.10] | 1.06  [1.00; 1.12] | 0.01  [-0.01; 0.04] | 0.01  [-0.01; 0.03] | 1.03  [0.97; 1.10] | 0.18*  [0.06; 0.30] | 1.06  [0.96; 1.17] |
|  | Distance to nearest blue space (SD) | 0.01  [-0.03; 0.04] | 1.00  [0.95; 1.06] | -0.01  [-0.04; 0.01] | -0.00  [-0.02; 0.01] | 0.96  [0.89; 1.02] | 0.01  [-0.10; 0.13] | 1.00  [0.90; 1.11] |

Abbreviations: BMI, body mass index; NDVI, normalized difference vegetation index; CI, confidence interval; SD, standard deviation * p-value < 0.05; β values are regression coefficients (95% confidence interval) from linear regression models, reflecting the change in body mass index z-score, systolic and diastolic blood pressure z-scores, and metabolic syndrome score, per unit of increase of each standard deviation of exposure, at each follow-up. OR values are odds ratios (95% confidence interval) from logistic regression models, reflecting the odds of having overweight/obesity, high blood pressure, and metabolic syndrome at 10 years, per unit of increase of each standard deviation of exposure, at each follow-up

**Table S4**. Crude and adjusted linear regression associations of green and blue spaces, at each time point, with continuous body fat content and fat distribution outcomes at 10 years (n=2623)

|  |  | **Fat mass index** | | **Android-to-gynoid fat ratio** | |
| --- | --- | --- | --- | --- | --- |
|  |  | **Crude model** | **Adjusted model** | **Crude model** | **Adjusted model** |
|  |  | **Exp(β) [95% CI]** | **Exp(β) [95% CI]** | **β [95% CI]** | **β [95% CI]** |
| **At birth** | NDVI 100m (SD) | 1.02  [1.01;1.04] | 1.02  [1.00;1.03] | 0.01  [0.00;0.01] | 0.01  [0.00;0.01] |
|  | NDVI 250 m (SD) | 1.02  [1.00; 1.03] | 1.01  [0.99;1.03] | 0.01  [0.00; 0.01] | 0.00  [-0.00;0.01] |
|  | NDVI 500m (SD) | 1.02  [1.00; 1.03] | 1.01  [0.99;1.03] | 0.01  [0.00;0.01] | -0.02  [-0.00;0.02] |
|  | Distance to nearest green space (SD) | 1.01  [1.00; 1.03] | 1.00  [0.99;1.02] | 0.00  [-0.00;0.01] | 0.00  [-0.01;0.01] |
|  | Distance to nearest blue space (SD) | 0.99  [0.98; 1.01] | 0.99  [0.97;1.00] | -0.00  [-0.01;0.00] | -0.00  [-0.01;0.00] |
| **4 years** | NDVI 100m (SD) | 1.01  [1.00; 1.03] | 1.01  [0.99;1.02] | 0.00  [-0.00; 0.01] | 0.00  [-0.01;0.01] |
|  | NDVI 250 m (SD) | 1.01  [1.00; 1.03] | 1.00  [0.98;1.02] | 0.00  [-0.00; 0.01] | 0.00  [-0.00;0.01] |
|  | NDVI 500m (SD) | 1.01  [1.00; 1.03] | 1.00  [0.98;1.02] | 0.00  [-0.00; 0.01] | 0.00  [-0.00;0.01] |
|  | Distance to nearest green space (SD) | 1.01  [1.00;1.03] | 1.00  [0.99;1.02] | 0.00  [-0.00; 0.01] | 0.00  [-0.01;0.01] |
|  | Distance to nearest blue space (SD) | 0.99  [0.97;1.00] | 0.98  [0.97;1.00] | -0.00  [-0.01; 0.01] | -0.00  [-0.01;0.01] |
| **7 years** | NDVI 100m (SD) | 1.01  [1.00; 1.03] | 1.01  [0.90;1.02] | 0.00  [-0.00; 0.01] | 0.00  [-0.00;0.01] |
|  | NDVI 250 m (SD) | 1.01  [1.00; 1.03] | 1.01  [1.00;1.08] | 0.01  [-0.00; 0.01] | 0.00  [-0.00;0.01] |
|  | NDVI 500m (SD) | 1.01  [1.00; 1.02] | 1.00  [0.98;1.02] | 0.01  [-0.00; 0.01] | 0.00  [-0.00;0.01] |
|  | Distance to nearest green space (SD) | 1.01  [1.00; 1.03] | 1.01  [0.99;1.02] | 0.00  [-0.00; 0.01] | 0.00  [-0.00;0.01] |
|  | Distance to nearest blue space (SD) | 0.99  [0.97; 1.00] | 0.98  [0.97;1.00] | -0.00  [-0.01;0.00] | -0.00  [-0.01;0.00] |
| **10 years** | NDVI 100m (SD) | 1.01  [0.99; 1.02] | 1.00  [0.98;1.02] | 0.00  [-0.00; 0.01] | 0.00  [-0.01;0.01] |
|  | NDVI 250 m (SD) | 1.00  [0.99; 1.02] | 1.00  [0.98;1.01] | 0.00  [-0.00; 0.01] | -0.00  [-0.01;0.01] |
|  | NDVI 500m (SD) | 1.00  [0.99; 1.02] | 0.99  [0.97;1.01] | 0.00  [-0.00; 0.01] | 0.00  [-0.01;0.01] |
|  | Distance to nearest green space (SD) | 1.01  [0.99; 1.02] | 1.00  [0.99;1.02] | 0.00  [-0.00; 0.01] | 0.00  [-0.01;0.01] |
|  | Distance to nearest blue space (SD) | 0.99  [0.98; 1.01] | 0.99  [0.97;1.00] | 0.00  [-0.01;0.01] | -0.00  [-0.01;0.01] |

Abbreviations: NDVI, normalized difference vegetation index; CI, confidence interval; SD, standard deviation. β and Exponential (Exp) β values are regression coefficients and their exponentiation (95% confidence interval) from linear regression models, reflecting the absolute change in android-to-gynoid fat ratio and the relative change in fat mass index, respectively, per unit of increase of each standard deviation of exposure, at each follow-up. Adjusted β and exp(β) consider maternal educational level at birth, and neighbourhood deprivation index, nitrogen dioxide (NO_2_) and particulate matter less than 2.5 μm (PM2.5) at each respective time point

**Table S5.** Criteria to assess model fit for latent group analysis model for Normalized Difference Vegetation Index (NDVI) and distance to the nearest green and blue space

|  | **Number of groups** | | | |
| --- | --- | --- | --- | --- |
|  | 1 | 2 | 3 | 4 |
| **NDVI 100m** |  |  |  |  |
| *AIC* | -102880.1 | -103636.2 | -103628.2 | -103620.2 |
| *BIC* | -102810.8 | **-103539.2** | -103503.5 | -103467.8 |
| **NDVI 250m** |  |  |  |  |
| *AIC* | -109390.3 | -109602.3 | -110668.3 | -112102.2 |
| *BIC* | -109321.0 | -109519.1 | -110543.6 | **-111949.7** |
| **NDVI 500m** |  |  |  |  |
| *AIC* | -115646.1 | -116213.4 | -117141.8 | -118397.6 |
| *BIC* | -115576.8 | -116116.4 | -117017.1 | **-118245.2** |
| **Distance to the nearest green space (hm)** |  |  |  |  |
| *AIC* | 38846.9 | 37239.1 | 33850.5 | 33858.4 |
| *BIC* | 38916.2 | 37336.1 | **33975.2** | 34011.0 |
| **Distance to the nearest blue space (hm)** |  |  |  |  |
| *AIC* | 33959.7 | 33280.8 | 33224.9 | 33296.8 |
| *BIC* | 34028.9 | **33377.9** | 33349.6 | 33449.3 |
| Abbreviations: NDVI, normalized difference vegetation index; AIC, Akaike Information Criterion; BIC, Bayesian Information Criterion; hm, hectometre | | | | |

**Table S6.** Probability of belonging to each latent class for Normalized Difference Vegetation Index (NDVI) and distance to the nearest green and blue space

|  | **Number of groups** | | | |
| --- | --- | --- | --- | --- |
|  | **1** | **2** | **3** | **4** |
| **NDVI 100m** |  |  |  |  |
| *Probability 1* | 0.8325 | 0.0528 |  |  |
| *Probability 2* | 0.1675 | 0.9472 |  |  |
| **NDVI 250m** |  |  |  |  |
| *Probability 1* | 0.8981 | 0.0061 | 0.0024 | 0.0000 |
| *Probability 2* | 0.0874 | 0.9186 | 0.1596 | 0.0660 |
| *Probability 3* | 0.0145 | 0.0719 | 0.8315 | 0.0425 |
| *Probability 4* | 0.0000 | 0.0033 | 0.0064 | 0.8915 |
| **NDVI 500m** |  |  |  |  |
| *Probability 1* | 0.8981 | 0.0000 | 0.0076 | 0.0028 |
| *Probability 2* | 0.0000 | 0.9028 | 0.0024 | 0.0052 |
| *Probability 3* | 0.0809 | 0.0484 | 0.9093 | 0.1378 |
| *Probability 4* | 0.0211 | 0.0488 | 0.0806 | 0.8542 |
| **Distance to the nearest green space (hm)** | | | | |
| *Probability 1* | 0.9976 | 0.0504 | 0.0368 |  |
| *Probability 2* | 0.0015 | 0.9496 | 0.0000 |  |
| *Probability 3* | 0.0010 | 0.0000 | 0.9632 |  |
| **Distance to the nearest blue space (hm)** | | | | |
| *Probability 1* | 0.9558 | 0.1692 |  |  |
| *Probability 2* | 0.0442 | 0.8308 |  |  |

Abbreviations: NDVI, normalized difference vegetation index; hm, hectometre

**Table S7**. Description of the natural spaces exposure trajectories

|  | **Trajectories** | **Sample used for deriving trajectories**  **(n=7549)**  **n (%)** | **Study sample**  **(n=4669)**  **n (%)** |
| --- | --- | --- | --- |
| **NDVI 100m** | Low ascending | 6334 (83.9) | 3922 (84.0) |
|  | High stable (ref) | 1215 (16.1) | 747 (16.0) |
| **NDVI 250m** | Low ascending | 5248 (69.5) | 3223 (69.0) |
|  | Descending | 233 (3.1) | 148 (3.2) |
|  | Ascending | 298 (3.9) | 207 (4.4) |
|  | High stable (ref) | 1770 (23.4) | 1091 (23.4) |
| **NDVI 500m** | Low ascending | 4914 (65.1) | 3007 (64.4) |
|  | Descending | 216 (2.9) | 148 (3.2) |
|  | Ascending | 332 (4.4) | 227 (4.9) |
|  | High stable (ref) | 2087 (27.6) | 1287 (27.6) |
| **Distance to nearest green space** | Medium stable (ref) | 7215 (95.6) | 4396 (94.2) |
|  | Descending | 161 (2.1) | 136 (2.9) |
|  | Ascending | 173 (2.3) | 137 (2.9) |
| **Distance to nearest blue space** | Low stable (ref) | 861 (11.4) | 528 (11.3) |
|  | High stable | 6688 (88.6) | 4141 (88.7) |

Abbreviations: NDVI, normalized difference vegetation index

**Table S8**. Crude linear and logistic regression associations between green and blue spaces trajectories with continuous and dichotomic cardiometabolic outcomes at 10 years

|  |  | **BMI**  **z-scores**  **(n=4666)** | **Overweight/**  **Obesity**  **(n=4618)** | **Systolic blood pressure**  **z-scores**  **(n=4658)** | **Diastolic blood pressure**  **z-scores**  **(n=4658)** | **High blood**  **pressure**  **(n=4658)** | **Metabolic**  **Syndrome**  **Score**  **(n=2731)** | **Metabolic**  **Syndrome**  **(n=2731)** |
| --- | --- | --- | --- | --- | --- | --- | --- | --- |
|  |  | **β [ CI 95%]** | **OR [95% CI]** | **β [ CI 95%]** | **β [ CI 95%]** | **OR [95% CI]** | **β [ CI 95%]** | **OR [95% CI]** |
| **NDVI 100m** | Low ascending | -0.05  [-0.14;0.05] | 1.00  [0.86; 1.18] | -0.04  [-0.11; -0.03] | -0.030  [-0.078; 0.018] | 0.87  [0.73; 1.04] | -0.21  [-0.51; 0.10] | 0.92  [0.71; 1.21] |
|  | High stable | Ref. | Ref. | Ref. | Ref. | Ref. | Ref. | Ref. |
| **NDVI 250m** | Low ascending | -0.05  [-0.13;0.04] | 0.99  [0.86; 1.14] | -0.01  [-0.07; 0.05] | -0.021  [-0.063; 0.021] | 1.01  [0.86; 1.18] | -0.15  [-0.41; 0.12] | 0.98  [0.78; 1.24] |
|  | Descending | -0.11  [-0.32;0.11] | 0.92  [0.64; 1.30] | -0.02  [-0.16; 0.13] | -0.077  [-0.182; 0.028] | 0.96  [0.64; 1.41] | -0.76*  [-1.44; -0.09] | 0.69  [0.34; 1.30] |
|  | Ascending | -0.08  [-0.26;0.11] | 1.01  [0.75; 1.37] | -0.05  [-0.18; 0.07] | -0.044  [-0.135; 0.047] | 0.98  [0.70; 1.37] | -0.18  [-0.77; 0.41] | 0.82  [0.46; 1.40] |
|  | High stable | Ref. | Ref. | Ref. | Ref. | Ref. | Ref. | Ref. |
| **NDVI 500m** | Low ascending | -0.09  [-0.17; -0.01] | 0.94  [0.82; 1.07] | 0.00  [-0.06; 0.06] | -0.004  [-0.044; 0.036] | 0.97  [0.84; 1.13] | -0.12  [-0.37; 0.13] | 1.00  [0.80; 1.25] |
|  | Descending | -0.23*  [-0.43; -0.02] | 0.84  [0.59; 1.18] | -0.02  [-0.16; 0.13] | -0.143*  [-0.247; -0.039] | 0.83  [0.55; 1.22] | -0.73*  [-1.35; -0.09] | 0.67  [0.34; 1.21] |
|  | Ascending | -0.09  [-0.26;0.09] | 1.07  [0.80; 1.42] | -0.10  [-0.23; 0.02] | -0.055  [-0.141; 0.032] | 0.76  [0.54; 1.05] | -0.38  [-0.96; 0.19] | 0.63  [0.33; 1.09] |
|  | High stable | Ref. | Ref. | Ref. | Ref. | Ref. | Ref. | Ref. |
| **Distance to nearest green space** | Medium stable | Ref. | Ref. | Ref. | Ref. | Ref. | Ref. | Ref. |
|  | Descending | -0.05  [-0.26;0.16] | 1.08  [0.76; 1.52] | 0.02  [-0.13; 0.16] | 0.013  [-0.091;0.118] | 1.08  [0.73; 1.56] | 0.28  [-0.36; 0.92] | 1.17  [0.66; 1.97] |
|  | Ascending | 0.02  [-0.19;0.23] | 0.99  [0.70; 1.39] | 0.02  [-0.13; 0.16] | 0.030  [-0.074;0.135] | 0.93  [0.62; 1.37] | -0.04  [-0.70; 0.62] | 0.76  [0.38; 1.39] |
| **Distance to nearest blue space** | Low stable | Ref. | Ref. | Ref. | Ref. | Ref. | Ref. | Ref. |
|  | High stable | -0.07  [-0.19;0.04] | 0.96  [0.80; 1.16] | -0.06  [-0.14;0.02] | -0.024  [-0.079;0.032] | 0.88  [0.72; 1.08] | -0.22  [-0.57; 0.12] | 0.91  [0.68; 1.24] |

Abbreviations: BMI, body mass index; NDVI, normalized difference vegetation index; CI, confidence interval; * p-value < 0.05. β values are regression coefficients (95% confidence interval) from linear regression models, reflecting the change in body mass index z-score, systolic and diastolic blood pressure z-scores, and metabolic syndrome score, for each trajectory as compared to the reference trajectory. OR values are odds ratios (95% confidence interval) from logistic regression models, reflecting the odds of having overweight/obesity, high blood pressure, and metabolic syndrome at 10 years, for each trajectory as compared to the reference trajectory

**Table S9**. Crude and adjusted linear regression associations of green and blue spaces trajectories with continuous body fat content and fat distribution outcomes at 10 years (n=2623)

|  |  | **Fat mass index** | | **Android-to-gynoid fat ratio** | |
| --- | --- | --- | --- | --- | --- |
|  |  | **Crude model** | **Adjusted model** | **Crude model** | **Adjusted model** |
|  |  | **Exp(β) [95% CI]** | **Exp(β) [95% CI]** | **β [95% CI]** | **β [95% CI]** |
| **NDVI 100m** | Low ascending | 0.96  [0.92; 0.99] | 0.97  [0.93;1.02] | -0.01  [-0.03; 0.00] | -0.01  [-0.02;0.01] |
|  | High stable | Ref. | Ref. | Ref. | Ref. |
| **NDVI 250m** | Low ascending | 0.96  [0.92; 0.99] | 0.98  [0.94;1.01] | 0.02  [-0.03; 0.00] | -0.01  [-0.03;0.01] |
|  | Descending | 0.95  [0.87; 1.04] | 0.97  [0.89;1.06] | -0.01  [-0.05; 0.02] | -0.01  [-0.04;0.02] |
|  | Ascending | 0.94  [0.87; 1.00] | 0.97  [0.90;1.05] | 0.04  [-0.06; -0.01] | -0.03  [-0.05;0.00] |
|  | High stable | Ref. | Ref. | Ref. | Ref. |
| **NDVI 500m** | Low ascending | 0.97  [0.94; 1.00] | 0.99  [0.95;1.03] | -0.01  [-0,03; -0.00] | **-**0.01  [-0.02;0.01] |
|  | Descending | 0.96  [0.89; 1.05] | 0.97  [0.89; 1.06] | -0.02  [-0.05; 0.01] | -0.02  [-0.06;0.01] |
|  | Ascending | 0.97  [0.90; 1.03] | 1.01  [0.94;1.09] | -0.02  [-0.05; 0.00] | -0.01  [-0.04;0.02] |
|  | High stable | Ref. | Ref. | Ref. | Ref. |
| **Distance to nearest green space** | Medium stable | Ref. | Ref. | Ref. | Ref. |
|  | Descending | 0.99  [0.91;1.07] | 0.99  [0.91;1.08] | -0.01  [-0.04; 0.03] | -0.00  [-0.04;0.03] |
|  | Ascending | 1.01  [0.93; 1.10] | 1.00  [0.92;1.09] | 0.00  [-0.03; 0.03] | -0.00  [-0.03;0.03] |
| **Distance to nearest blue space** | Low stable | Ref. | Ref. | Ref. | Ref. |
|  | High stable | 0.98  [0.94; 1.03] | 0.98  [0.94;1.03] | -0.00  [-0.02; 0.02] | -0.00  [-0.02;0.02] |

Abbreviations: NDVI, normalized difference vegetation index; CI, confidence interval; β and Exponential (Exp) β values are regression coefficients and their exponentiation (95% confidence interval) from linear regression models, reflecting the absolute change in android-gynoid index and the relative change in fat mass index, respectively, for each trajectory as compared to the reference trajectory. Adjusted β and exp(β) consider maternal educational level at birth, and neighbourhood deprivation index, nitrogen dioxide (NO_2_) and particulate matter less than 2.5 μm (PM2.5) at birth and at 10 years

**­­­­­­­ Table S10**. Adjusted linear regression associations of green and blue spaces, at each time point, with continuous z-scores of metabolic syndrome components at 10 years (n=2731)

|  | | **Waist circunference**  **z-scores** | **Triglycerides**  **z-scores** | **HDL**  **z-scores** | **HOMA-IR**  **z-scores** | **Glucose**  **z-scores** |
| --- | --- | --- | --- | --- | --- | --- |
|  |  | **β [95% CI]** | | | | |
| **At birth** | NDVI 100m (SD) | 0.03  [-0.03;0.08] | -0.02  [-0.05;0.01] | 0.01  [-0.03;0.04] | 0.02  [-0.03;0.07] | 0.02  [-0.01; 0.05] |
|  | NDVI 250 m (SD) | 0.01  [-0.06;0.07] | -0.02  [-0.05;0.02] | -0.01  [-0.05;0.03] | -0.01  [-0.07;0.05] | 0.00  [-0.04; 0.04] |
|  | NDVI 500m (SD) | -0.01  [-0.08;0.07] | -0.03  [-0.07;0.01] | 0.01  [-0.03;0.05] | -0.03  [-0.10;0.03] | -0.02  [-0.06; 0.02] |
|  | Distance to nearest green space (SD) | 0.02  [-0.04;0.08] | 0.01  [-0.01;0.04] | -0.03  [-0.06; 0.00] | 0.04  [-0.01;0.09] | 0.01  [-0.02; 0.04] |
|  | Distance to nearest blue space (SD) | 0.03  [-0.03;0.08] | -0.01  [-0.03;0.02] | -0.01  [-0.04;0.02] | -0.02  [-0.07;0.02] | -0.03*^†^  [-0.06; -0.00] |
| **4 years** | NDVI 100m (SD) | 0.02  [-0.04;0.08] | -0.02  [-0.05;0.01] | 0.00  [-0.03;0.04] | 0.02  [-0.03;0.07] | 0.03  [-0.00; 0.06] |
|  | NDVI 250 m (SD) | 0.03  [-0.03;0.10] | 0.01  [-0.04;0.03] | -0.02  [-0.06;0.02] | 0.02  [-0.04;0.08] | 0.01  [-0.02; 0.05] |
|  | NDVI 500m (SD) | 0.03  [-0.04;0.10] | -0.02  [-0.06;0.02] | 0.00  [-0.04;0.05] | -0.00  [-0.06; 0.06] | -0.01  [-0.04; 0.03] |
|  | Distance to nearest green space (SD) | 0.05  [-0.01;0.10] | 0.01  [-0.02;0.04] | -0.02  [-0.05;0.02] | 0.06*^†^  [0.01;0.11] | 0.02  [-0.02; 0.05] |
|  | Distance to nearest blue space (SD) | 0.04  [-0.01;0.09] | -0.02  [-0.04;0.01] | -0.01  [-0.04;0.02] | -0.02  [-0.07;0.02] | -0.04*^†^  [-0.06; -0.01] |
| **7 years** | NDVI 100m (SD) | 0.03  [-0.03;0.09] | -0.01  [-0.04;0.02] | 0.00  [-0.03;0.03] | -0.01  [-0.07;0.04] | 0.01  [-0.02;0.05] |
|  | NDVI 250 m (SD) | 0.03  [-0.03;0.10] | 0.00  [-0.03;0.04] | -0.02  [-0.06;0.02] | -0.02  [-0.07;0.04] | -0.00  [-0.04; 0.03] |
|  | NDVI 500m (SD) | 0.02  [-0.05;0.09] | -0.01  [-0.05;0.02] | -0.00  [-0.05;0.04] | -0.04  [-0.10;0.03] | -0.03  [-0.07;0.01] |
|  | Distance to nearest green space (SD) | 0.06  [-0.00;0.11] | 0.01  [-0.01;0.04] | -0.02  [-0.06;0.01] | 0.05  [-0.00;0.10] | 0.02  [-0.01;0.05] |
|  | Distance to nearest blue space (SD) | 0.04  [-0.01;0.09] | -0.03*^†^  [-0.05; -0.00] | -0.00  [-0.03;0.03] | -0.03  [-0.08;0.02] | -0.04*^†^  [-0.07; -0.01] |
| **10 years** | NDVI 100m (SD) | 0.02  [-0.04;0.08] | 0.00  [-0.03;0.03] | -0.02  [-0.05;0.02] | 0.02  [-0.03;0.08] | 0.02  [-0.02;0.05] |
|  | NDVI 250 m (SD) | 0.02  [-0.05;0.09] | 0.02  [-0.02;0.05] | -0.04*^†^  [-0.08; -0.00] | 0.04  [-0.02; 0.10] | 0.02  [-0.01;0.06] |
|  | NDVI 500m (SD) | -0.01  [-0.07;0.06] | -0.00  [-0.04;0.03] | -0.01  [-0.05;0.03] | 0.02  [-0.04;0.08] | -0.02  [-0.06; 0.02] |
|  | Distance to nearest green space (SD) | 0.05  [-0.00;0.11] | 0.02  [-0.01;0.05] | -0.02  [-0.06;0.01] | 0.05  [0.00;0.10] | 0.01  [-0.02;0.04] |
|  | Distance to nearest blue space (SD) | 0.03  [-0.02; 0.08] | -0.03  [-0.05;0.00] | -0.01  [-0.04;0.02] | -0.01  [-0.06;0.03] | -0.04*^†^  [-0.07; -0.01] |

Abbreviations: NDVI, normalized difference vegetation index; CI, confidence interval; SD, standard deviation . * p-value < 0.05; ^†^ Indicates loss of significance after multiple testing correction. β values are regression coefficients (95% confidence interval) from linear regression models, reflecting the change in waist circumference, triglycerides, HDL, HOMA-IR, and glucose z-scores, per unit of increase of each standard deviation of exposure, at each follow-up. Adjusted β consider maternal educational level at birth, and neighbourhood deprivation index, nitrogen dioxide (NO2) and particulate matter less than 2.5 μm (PM2.5) at each respective timepoint

**Table S11**. Adjusted linear regression associations between green and blue spaces trajectories with continuous z-scores of metabolic syndrome components at 10 years (n=2731)

|  |  | **Waist Circumference**  **z-scores** | **Triglycerides**  **z-scores** | **HDL**  **z-scores** | **HOMA-IR**  **z-scores** | **Glucose**  **z-scores** |
| --- | --- | --- | --- | --- | --- | --- |
|  |  | **β [95% CI]** | | | | |
| **NDVI 100m** | Low ascending | 0.01  [-0.15;0.16] | 0.01  [-0.07;0.08] | 0.01  [-0.08;0.10] | -0.02  [-0.16;0.12] | -0.02  [-0.11;0.06] |
|  | High stable | Ref. | Ref. | Ref. | Ref. | Ref. |
| **NDVI 250m** | Low ascending | 0.01  [-0.14;0.15] | 0.03  [-0.05;0.10] | 0.05  [-0.03;0.13] | 0.01  [-0.12;0.14] | 0.03  [-0.05;0.11] |
|  | Descending | -0.29  [-0.60;0.02] | -0.16  [-0.05; 0.10] | 0.13  [-0.06;0.31] | -0.13  [-0.41;0.16] | 0.01  [-0.17;0.19] |
|  | Ascending | -0.00  [-0.28;0.28] | 0.10  [-0.05;0.24] | 0.04  [-0.13;0.21] | 0.00  [-0.25;0.26] | -0.01  [-0.17;0.16] |
|  | High stable | Ref. | Ref. | Ref. | Ref. | Ref. |
| **NDVI 500m** | Low ascending | -0.00  [-0.14;0.14] | 0.02  [-0.05;0.09] | 0.05  [-0.03;0.13] | 0.03  [-0.09;0.16] | 0.05  [-0.03;0.12] |
|  | Descending | -0.31*^†^  [-0.61;-0.01] | -0.03  [-0.19;0.12] | -0.05  [-0.13;0.22] | -0.18  [-0.45;0.09] | -0.01  [-0.18;0.16] |
|  | Ascending | 0.03  [-0.25;0.31] | 0.00  [-0.14;0.15] | 0.01  [-0.15;0.18] | -0.11  [-0.36;0.14] | -0.08  [-0.23;0.08] |
|  | High stable | Ref. | Ref. | Ref. | Ref. | Ref. |
| **Distance to nearest green space** | Medium stable | Ref. | Ref. | Ref. | Ref. | Ref. |
|  | Descending | 0.03  [-0.28;0.33] | 0.05  [-0.11;0.20] | 0.04  [-0.13;0.22] | -0.05  [-0.32;0.22] | -0.07  [-0.24;0.10] |
|  | Ascending | 0.00  [-0.29;0.29] | 0.08  [-0.07;0.23] | -0.02  [-0.20;0.15] | 0.10  [-0.16;0.37] | -0.01  [-0.17;0.16] |
| **Distance to nearest blue space** | Low stable | Ref. | Ref. | Ref. | Ref. | Ref. |
|  | High stable | -0.03  [-0.19;0.12] | -0.04  [-0.13;0.04] | -0.04  [-0.13;0.06] | -0.14  [-0.28;0.01] | -0.10*^†^  [-0.19; -0.01] |

Abbreviations: NDVI, normalized difference vegetation index. CI, confidence interval. * p-value < 0.05; ^†^ Indicates loss of significance after multiple testing correction. β values are regression coefficients (95% confidence interval) from linear regression models, reflecting the change in waist circumference, triglycerides, HDL, and glucose z-scores, for each trajectory as compared to the reference trajectory. Adjusted β consider maternal educational level at birth, and neighbourhood deprivation index, nitrogen dioxide (NO_2_) and particulate matter less than 2.5 μm (PM2.5) at birth and at 10 years
